# Early response model for containing newly emerging infectious diseases

**DOI:** 10.1101/2025.10.14.25337721

**Authors:** Marcus P.S. Dekens, Tobias Neumann, Thomas Micheler, Vinika Gandhi, Max J. Kellner, Marianne Rocha-Hasler, Martina Weißenböck, Michaela Fellner, Mariam Al-Rawi, Nikolaus Beer, Sarah Rieser, Christian Umkehrer, David Hoffmann, Martin Mátl, Katrin Künstner, Vienna COVID-19 Detection Initiative, Zlatka Cular, Sabine Walch, Johanna Trupke, Peter Steinlein, Harald Scheuch, Thomas Wochele-Thoma, Robert Heinen, Johannes Zuber

## Abstract

**Background:** Recent epidemics of influenza, AIDS, SARS, Ebola, Zika, and COVID-19 highlight the persistent threat of RNA viruses to public health. COVID-19 exposed major gaps in monitoring capabilities, leaving lockdowns as the only means to reduce transmission. Moreover, their effectiveness was limited by economic pressures that forced early relaxation.

**Methods:** To strengthen monitoring capabilities, we designed a scalable workflow for specimen collection, analysis, and reporting. Rapid implementation was achieved by repurposing non medical, nonprofit, laboratory infrastructure. We analyzed the variation in 4,295,664 viral genomes collected from patients worldwide using software developed inhouse. qPCR assays targeting stable regions of the pathogen’s genome were developed and then validated in a clinical performance study. Between September 2020 and December 2021, 476,502 specimens from local staff and caregivers were analyzed, 177,756 of which were processed in batches.

**Results:** The effectiveness of this response model is demonstrated by the significant reduction in incidence (*P*=0.007, *d*=4.54) among 2,475 systematically monitored caregivers compared with the general population. Performance validation of the assays showed high analytical sensitivity, enabling scaling up while detecting SARS-CoV-2 below the infectious viral load. Frequent testing with same-day notification was associated with effective control of viral transmission, comparing favorably with lockdowns. The high inclusive specificity (>99.99%) of the diagnostic panel further minimized outbreak risk.

**Interpretation:** Timely identification and isolation of cases is essential to protect health systems, inform policy decisions, and support economic stability. Infectious diseases with pre- or asymptomatic transmission necessitate molecular diagnostic testing. Effective deployment requires dedicated workflows that integrate supply and information systems, combining high-throughput with fast, affordable results. This after-action report presents an operationally grounded framework for responding to emerging infectious diseases.

## Introduction

An infectious disease outbreak occurs when there is little or no pre-existing immunity in the population, which can lead to an epidemic and spread globally if not quickly contained. Pathogens that can spread fast through human populations have traits that make them hard to avoid, such as airborne, presymptomatic and asymptomatic transmission. Vaccination campaigns are the most effective defense against infectious diseases, as vaccines prepare the immune system to fight against them, reducing disease severity, hospitalizations and mortality. However, developing, mass producing and distributing vaccines takes time. Therefore, the most effective early response to protect public health and maintain economic stability is the rapid identification and isolation of infected individuals.

Testing also provides valuable information that informs decision-making. Thus, the reliability of outbreak assessments depends on the extent of testing. Insufficient testing masks true case numbers, leading to delayed interventions and premature relaxation of measures. This allows residual cases to initiate a new wave of infections [1, 2]. Delays provide the pathogen with time to accumulate mutations, followed by the selection of variants with enhanced infectivity and partial immune evasion. These variants of concern (VoC) increase the likelihood of subsequent infection surges [3]. Thus, to effectively control transmission, monitoring must begin immediately after the first cases are reported and achieve high population coverage.

Infectious disease monitoring differs from routine clinical laboratory operations in that its primary goal is to control the transmission of a pathogen rather than individual patient care. This requires a shift in the diagnostic process. For monitoring, testing frequency and turnaround time are more critical than analytical sensitivity [4-6]. As it is sufficient to identify infected individuals before they become contagious, capacity can be expanded through batch testing when using a highly sensitive method [7-9]. The method’s specificity must be both exclusive and inclusive. Exclusive specificity ensures that uninfected individuals test negative. Pathogens with epidemic potential tend to be RNA viruses that mutate at high rates [10]. Variant-inclusive specificity prevents emerging variants from evading detection, thereby minimizing outbreak risk. Quantitative polymerase chain reaction (qPCR) is the preferred method because it combines high sensitivity and specificity with broad pathogen detectability. By contrast, rapid antigen tests, while fast and easy to use, have lower sensitivity, missing individuals in the early contagious stage [11]. Moreover, following an infectious disease outbreak, qPCR assays can be developed in one month due to their modular design, whereas antigen assays require time-consuming antibody production. Since testing frequency is essential, purpose-built high-throughput workflows with integrated supply and information systems are as important as the diagnostic assay itself.

On December 31, 2019, the World Health Organization (WHO) country office reported an outbreak of an atypical pneumonia-like illness of unknown etiology in Wuhan, China. On March 11, 2020, the WHO declared the identified coronavirus disease (COVID-19) a pandemic. COVID is caused by severe acute respiratory syndrome coronavirus 2 (SARS-CoV-2), a single-stranded positive-sense RNA virus [12, 13]. The infection begins with rapid replication in the nasopharynx and may progress to acute respiratory failure [14]. The combination of airborne with pre- and asymptomatic transmission made SARS-CoV-2 particularly difficult to control [15-18]. Because most governments lacked the core capabilities for rapid detection and response, the only immediate option to avert health system collapse was to slow transmission through lockdowns. While effective in reducing critically ill patients, the lockdowns had to be cut short as they brought the economy to a standstill.

During the pandemic, our mission was to establish high-throughput cost-effective monitoring, validate this approach as a means of mitigating virus transmission, and support vulnerable groups. Implementation requires infrastructure consisting of specialist staff, resources, safe workspaces and information systems. To rapidly establish monitoring, we repurposed the nonprofit infrastructure of the Vienna BioCenter (VBC). To minimize outbreak risk, we developed variant-inclusive qPCR assays targeting stable regions of the SARS-CoV-2 genome, identified through analysis of genomes available through GISAID (Global Initiative on Sharing All Influenza Data) [19, 20]. Over one year, we monitored the staff of research institutes and caregivers at 17 care homes, conducting 476,502 tests. To contain the spread of newly emerging infectious diseases, a scalable monitoring model is needed that can be rapidly deployed and protects public health and the economy until a vaccine becomes available. Here, we present a validated blueprint for infectious disease monitoring as an integral part of a response strategy.

## Results

An operational plan was created to establish an infectious disease monitoring system including specimen collection, diagnostic analysis and reporting (Figure S1A-D). With laboratories and expertise in place, RT-qPCR assays were rapidly developed. The published target sequences of the SARS-CoV-2 genome from the Charité [21] and the U.S. Centers for Disease Control and Prevention (CDC) [22] obviated the need for oligonucleotide design, although performance testing remained necessary. The use of commercially available reagents further accelerated development. The in-house assays allowed us to overcome the global shortage of diagnostic kits, rapidly adapt to emerging variants, and reduce costs. A major challenge in monitoring is the labor-intensive collection of nasopharyngeal swabs at centralized stations. To improve compliance with regular testing and reduce costs, we introduced self-sampling by gargling. This method has been proven reliable for virus detection [23] and was validated in our system. On campus, all specimens were deposited at a central outdoor collection point, while monitoring of caregivers required more advanced logistics because the care homes are dispersed across the city.

We first implemented a small-scale workflow with a daily testing capacity of ∼500 specimens, with VBC employees volunteering to participate in anonymous monitoring. This allowed optimization of the workflow before scaling up. Capacity was then increased through the establishment of a facility. High-throughput, error-free pipetting, and reproducibility were achieved by automating all stages of the workflow (Figure 1). To prevent amplicon contamination the stages were separated into clean, low-copy, and high-copy rooms (Figure S2). To expedite specimen handling, ensure data integrity, and deliver results directly to participants, we developed a laboratory information management system application compliant with the General Data Protection Regulation (Figure S1B). To further expand capacity, we introduced two-stage batch testing: pooling specimens for initial testing, followed by testing individual specimens from positive batches. The batch testing included nucleic acid extraction to maximize the sensitivity of the RT-qPCR. While batch testing increases capacity and reduces reagent costs, the deconvolution of positive batches has the opposite effect. In this case, a more efficient approach, performing RT-qPCR directly with each crude backup specimen eliminates extraction costs and shortens turnaround time. Direct RT-qPCR requires inactivation of RNases and viruses in the specimens. To achieve this, we validated an existing inactivation method using the reducing agent TCEP combined with heat [24], and selected assays that only amplify viral RNA from throat wash containing both RNA and genomic DNA.

**Figure 1.**
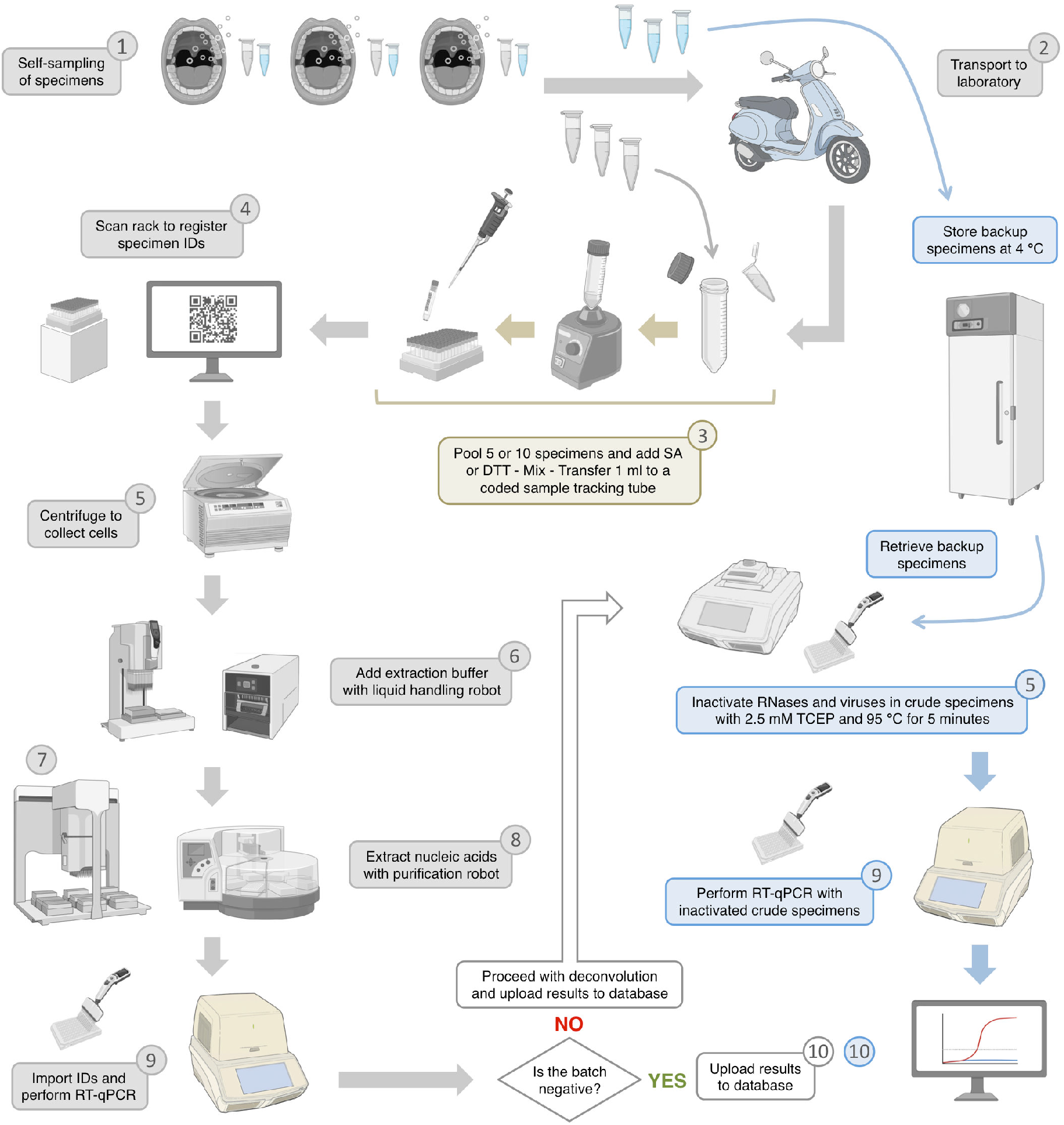
Scalable workflow for infectious disease monitoring. Specimens are self-sampled (1) and deposited in a dropbox, which is delivered to the laboratory (2). To scale up the workflow, batch testing was introduced, requiring two specimens per person: one specimen is pooled, and the other serves as a backup if the pool tests positive. Five or ten specimens are pooled and mixed with sodium ascorbate or DTT to reduce viscosity (3). The batch is then registered (4), centrifuged (5), and mixed with extraction buffer (6). Plate preparation (7), and nucleic acid extraction using robots (8), is followed by RT-qPCR (9). A qPCR machine can analyze 90 batches in one hour, yielding results from 450 or 900 specimens. To identify positive specimens within positive pools, RT-qPCR is performed directly on each crude backup specimen (blue arrows), reducing turnaround time and cost. After quality control, test results are uploaded to the database (10). To prevent amplicon contamination, workflow steps are spatially separated, as indicated by the numbers on the floor plan in figure S2.

On January 8, 2021, the U.S. Food and Drug Administration (FDA) warned that several authorized nucleic acid amplification test (NAAT) kits could potentially produce false negative results for new SARS-CoV-2 variants. Commercial kits do not disclose their target sequences, preventing independent validation. At the same time, mismatches between published assays and variants were reported [25]. With millions of SARS-CoV-2 genome sequences available through GISAID, we evaluated a set of widely used assays: E-Sarbeco, RdRP-SARSr [21], CDC-N1, CDC-N2, CDC-N3 [22], CCDC-N [26], CCDC-ORF1ab [27], and NIID-N [28]. In early 2021, we aligned the assays with ∼350,000 complete, high-coverage viral genomes collected from patients globally, and by December 2022 our final iteration included 4,295,664 genomes. To assess inclusive specificity, we defined a match as an exact sequence match between a viral genome and the assay. A high match rate corresponds to a low probability of false negatives. The RdRP-SARSr, CDC-N1 and CCDC-N assays showed strongly reduced match rates (Table 1, Figure 2A,C,D). To gain insight into the present mutational landscape, we analyzed 42,874 genomes collected at the end of our timeframe. E-Sarbeco, CDC-N1 and CCDC-N showed decreased match rates, while that of RdRP-SARSr increased, reflecting replacement of one virus lineage by another (Figure 2E-G, S3A,B). To reduce outbreak risk, assays that target different genomic regions were combined [29].

**Table 1.**
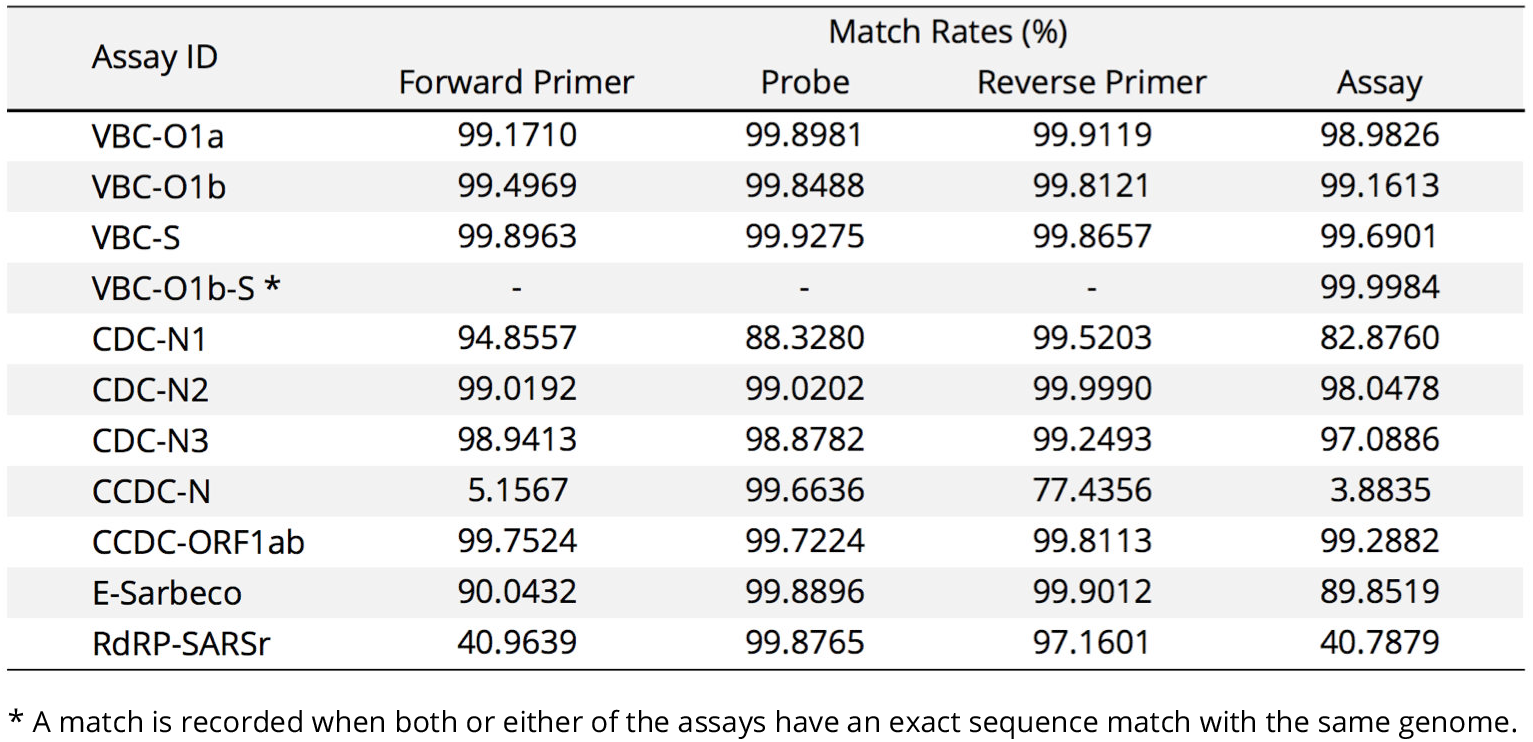
Match rates of assays based on 4,295,664 SARS-CoV-2 genomes.

**Figure 2.**
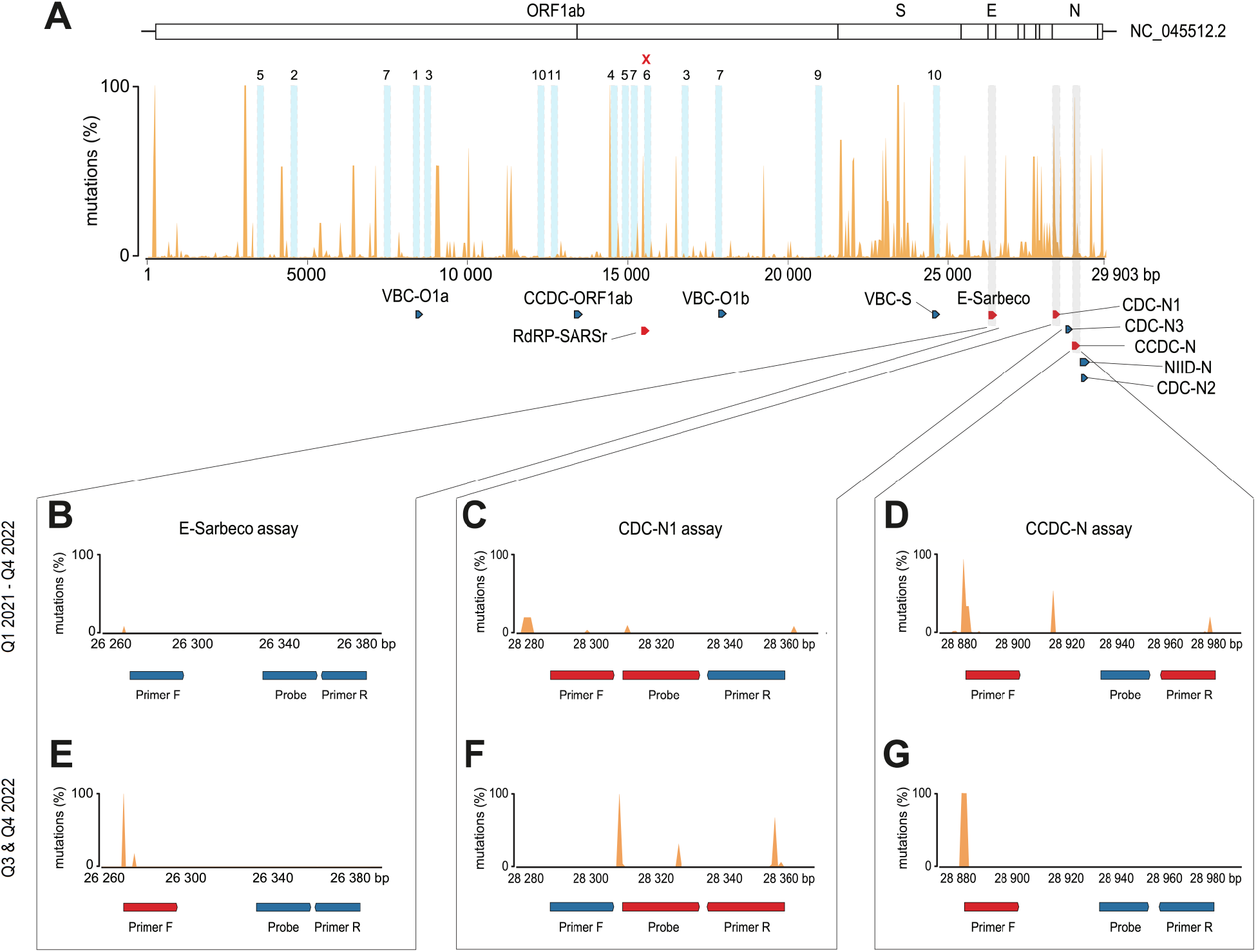
Cumulative mutation map of the SARS-CoV-2 genome. **A**. Mutation rate at each position in the SARS-CoV-2 reference genome, determined by aligning it with 4,295,664 complete, high6 coverage genomes collected from patients worldwide during Q1 2021 - Q4 2022. White bars above the map indicate gene locations. Vertical blue bars highlight stable regions, with the number of assays designed for each region shown above. Iterative analysis revealed that the region marked with a red cross became unstable over time and was therefore excluded. Bars below the map show assay target sites, with red bars indicating high mismatch rates. **B**. Mutation rate in the target region of the E-Sarbeco assay (Charité). Primer and probe target sites are shown below. **C**. As in B, for the US CDC-N1 assay. **D**. As in B, for the Chinese CDC-N1 assay. **E**. Mutation rate in the target region of the E6Sarbeco assay using 42,874 genomes collected in Q3-Q4 2022. **F**. As in E, for the US CDC-N1 assay. **G**. As in E, for the Chinese CDC-N assay.

To improve reliability for detecting circulating and newly emerging variants, we focused on stable regions of the viral genome. A cumulative mutation map of the SARS-CoV-2 genome was created by extracting mutations at each position in the reference sequence from alignments with 4,295,664 complete high-coverage viral genomes collected from patients globally (Figure 2A) [12]. Ninety candidate assays were designed that target stable regions within the SARS-CoV-2 genome (Figure S1C). Exclusive specificity was empirically validated against the oral microbiome, and by sequence comparison against SARS-CoV, the closest Sarbecovirus relative of SARS-CoV-2 that is capable of infecting humans [30]. As the VBC assays display multiple mismatches with SARS-CoV, its detection is highly unlikely (Table S2). Three assays targeting the viral replicase (ORF1) and spike (S) genes, as well as human β2-microglobulin RNA (B2M, control), were selected and subsequently compared to CDC-N2 and B2M in a clinical performance study (Table S1) [31].

The CDC-N2 assay was chosen as the reference due to its superior performance relative to other published assays [32]. Compared to CDC-N2 (98.05%), the VBC assays showed higher match rates: 99.16% for VBC-O1b and 99.69% for VBC-S, combined they exceeded 99.99% (Table 1). In addition to comparing assay match rates using a large variant pool, we demonstrate high variant-inclusive specificity for the VBC assays (Table S3).

The limit of detection (LoD) indicates whether the assay can detect low copy numbers of the virus. The LoD is defined as the copy number detected in 95% of replicates [33]. Throat wash from healthy individuals was spiked with a dilution series of AccuPlex standard: packaged complete viral genomes. These were purified followed by replicate RT-qPCR. LoDs were 5.7 copies/reaction for CDC-N2, 6.6 for VBC-O1b, and 5.7 for VBC-S (Figure 3A). The LoD for CDC-N2 is consistent with that reported by the CDC (5 copies/reaction) even though we included nucleic acid extraction from a matrix and used a different RT-qPCR kit [22]. As viral load correlates with transmissibility [34], the LoDs show that the VBC assays can detect SARS-CoV-2 well before an infected person becomes contagious, even when batch testing is used.

**Figure 3.**
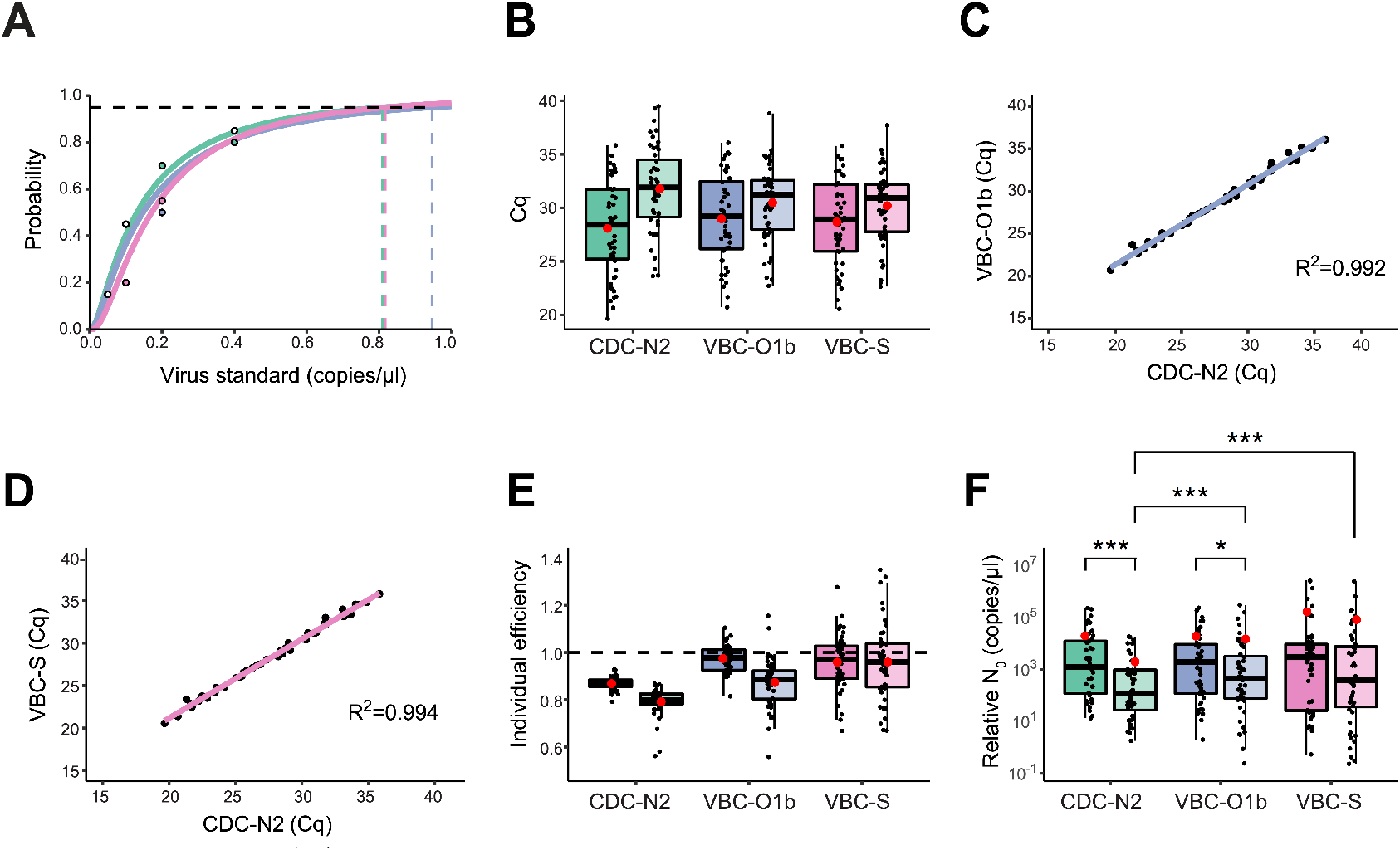
qPCR performance parameters of the CDC and VBC SARS-CoV-2 assays. **A**. Dilution PCR demonstrates high analytical sensitivity of the reference and VBC assays: CDC-N2 (0.81 copy/n μl, green line), VBC-O1b (0.95 copy/ μl, blue line) and VBC-S (0.82 copy/ μl, pink line). White dots indicate overlapping values. **B**. Boxplot of C_q_ values from 48 clinical specimens analyzed with CDC-N2, VBC-O1b, and VBC-S assays. Dark boxes represent qPCR with purified specimens; light boxes represent qPCR with crude specimens. Means are indicated by red dots. **C**. Accuracy of the VBC-O1b assay, showing that 99.2% of variation in test results from purified clinical specimens is shared with the reference (*P*=3.15×10^−50^). **D**. As in C, for the VBC-S assay, showing 99.4% agreement (*P*=4.55×10^−52^). **E** . Individual qPCR efficiencies calculated from fluorescence curves of reactions with viral RNA in each specimen. **F**. Relative initial copy number (N_0_) for each specimen tested across all assays. Asterisks indicate significant differences, bottom-left to top-right: *P*=3.44×10^−7^, *P*=0.02, *P*=6.39×10^−6^, *P*=3.65×10^−5^.

Reproducibility, assessed with the coefficient of variation in technical replicates, was high across assays for purified specimens, with CDC-N2 outperforming the VBC assays (Table S4). RT-qPCR with crude specimens showed reduced reproducibility across all assays.

In a non-interventional, non-randomized, retro-spective, open-label, single-laboratory clinical performance study, we compared the VBC-O1b-S-B2M with the CDC-N2-B2M reference using 48 clinical specimens. Each specimen was split for parallel testing of purified versus crude samples. CDC-N2 detected virus in purified samples an average of three cycles earlier than in crude samples, while the VBC assays showed only a cycle difference (Figure 3B, S4B). Agreement between the C_q_-values from the assays was measured using the coefficient of determination (R^2^). For purified specimens the agreement between the multiplexed VBC assays and CDC-N2 exceeded 99%, indicating high accuracy (Figure 3C,D, S4).

PCR efficiency, a measure of optimal amplification, was calculated from the fluorescence curve of each reaction with viral RNA in the clinical specimens using LinRegPCR software [35, 36]. The efficiency of VBC-O1b (*E*=97.4±1.6%, *P*=2.37×10^-14^) and VBC-S (*E*=95.9±3.2%, *P*=2.13×10^-6^) is significantly higher than that of CDC-N2 (*E*=86.8±0.8%) for purified specimens (Figure 3E). The reported higher efficiency of 99.4% for CDC-N2 may be due to measurements based on a standard curve instead of actual conditions [22]. With crude specimens, efficiencies were also significantly higher for the VBC-O1b (*E*=87.3±2.9%, *P*=1.07×10^-5^) and VBC-S (*E*=96.0±9.5%, *P*=7.66×10^-8^) compared with CDC-N2 (*E*=79.1±1.7%) (Figure 3E). While crude specimens reduced efficiency for all assays, VBC assays remained more robust (Table S5).

To assess differences in quantitative performance, relative initial copy numbers (N_0_) were determined (Figure 3F). This was calculated by applying a constant to the amplicon number (N_n_) in the amplification equation. No significant difference in relative N_0_ values was observed between CDC-N2, VBC-O1b and VBC-S for purified specimens. For crude specimens, VBC-O1b (*P*=6.39×10^-6^) and VBC-S (*P*=3.65×10^-5^) yielded higher relative N_0_ values.

During the pandemic, detection failure was the primary quality issue reported for NAAT testing, likely due to reduced assay design and processing standards [37]. In addition, emerging mutants that evade detection detection posed a major challenge for laboratories. Analysis of 4,295,664 million SARS-CoV-2 genomes revealed higher mutation rates at the 3’ end, which is targeted by the commonly used assays. To reduce outbreak risk, we developed qPCR assays that target stable genomic regions. Furthermore, the CDC-N2 and VBC assays showed high analytical sensitivity, enabling expansion of capacity through batch testing while detecting SARS-CoV-2 below the infectious viral load.

The second and third COVID surges occurred during the final quarter of 2020 and the first quarter of 2021. Systematic monitoring of caregivers started at the beginning of the second surge. Scaling was achieved by testing ten specimens in a batch. Monitoring resulted in a rapid decline of the weekly incidence rate from 1.35% to 0.60% prior to a city-wide hard lockdown (Figure 4A, Table S6). Two additional hard lockdowns followed, separated by a two-month period of relaxation. During this intervening period, the general population experienced a significant rise in new cases (*P*=0.012, *d*=2.52). However, no increase in incidence was observed among systematically monitored caregivers. The difference in incidence between monitored caregivers and the general population during the four weeks (10-13) before the third lockdown is significant (*P*=0.007, *d*=4.54). To further assess whether monitoring had reduced the incidence, we compared the four weeks (43-46) before the first lockdown with the four weeks (10-13) before the third. A significant reduction in incidence among monitored caregivers was observed (*P*=0.008, *d*=4.16), whereas the difference in incidence among the general population was not significant. To evaluate the potential impact of the third lockdown on incidence, we compared the four weeks (10-13) before the lockdown with the four weeks (18-21) after. The incidence among the general population showed a significant reduction (*P*=0.009, *d*=4.20), while no significant change was detected among the monitored caregivers. These results imply that city-wide hard lockdowns did not confer additional benefit beyond systematic monitoring, and that the initial reduction in incidence among caregivers during the first lockdown (43-49) was solely attributable to the monitoring. Notably, a transient peak in the incidence rate among caregivers occurred during the second lockdown (1-2), coinciding with the post festive season return of caregivers. Testing upon arrival and prompt isolation of positive staff resulted in a rapid decline in incidence, thereby preventing further transmission.

**Figure 4.**
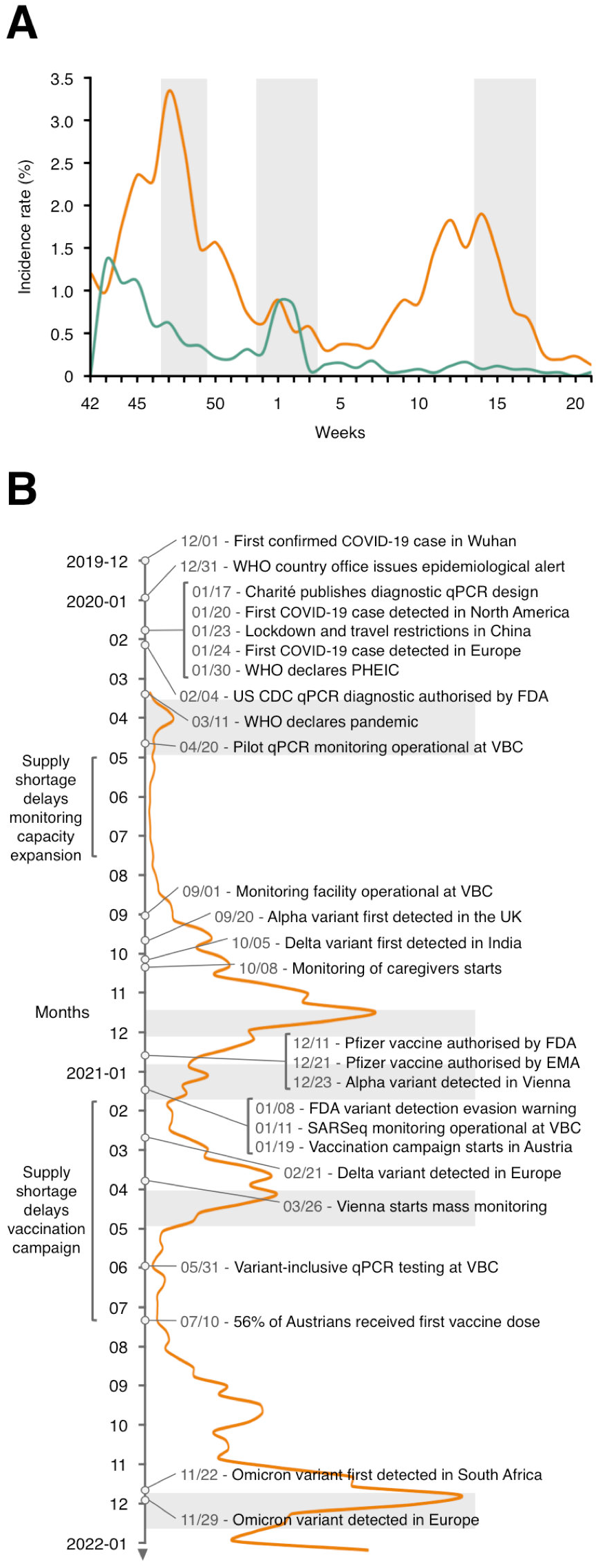
Comparison of monitoring with lockdowns: Improving risk mitigation. **A**. The weekly incidence rate of caregivers (green curve) compared with the population of Vienna (orange curve) during the second and third COVID waves shows a significant reduction in the incidence among systematically monitored caregivers. Hard lockdowns are highlighted by grey bars. The incidence in the general population is based on a relatively small percentage of individuals, whereas we tested caregivers systematically. City-wide lockdowns did not significantly reduce the incidence among monitored caregivers. **B**. Timeline of weekly incidence in Vienna, with alerts, measures, and variants of concern. Two months after the first confirmed case, the WHO declared a public health emergency of international concern (PHEIC). By that time, the virus had already spread from Wuhan in East Asia to Europe and North America. In the absence of a monitoring system, transmission could only be slowed through lockdowns. Economic pressure necessitated early relaxation of the lockdowns, which reduced their effectiveness. The Vienna BioCenter established small-scale monitoring within five weeks, but supply shortages delayed scaling capacity. The first vaccine was developed within nine months, but mass production and supply chain disruptions delayed vaccination campaigns.

## Discussion

The pandemic showed that prolonged and widespread infections become unavoidable when positive cases cannot be quickly identified and isolated. In the absence of an effective coordinated response, bottom-up initiatives emerged to create safe environments through monitoring. Because no such systems were in place, it became necessary to quickly establish an entire infrastructure, including trained staff, adequate resources, safe workspaces, and reliable information systems. Successful implementation depends on a well-defined framework and iterative improvement (Figure S1). Under optimal conditions, establishing monitoring takes at least three months, but delays are inevitable during an epidemic (Figure 4B). We established monitoring within five months by reallocating hard and soft resources from non medical nonprofit laboratories. We separated assay development from monitoring because the former requires flexibility that could compromise processing quality. Comparing the incidence rates of 2,475 systematically monitored caregivers with those of the general population demonstrates that a model based on frequent testing, same-day notification and contact tracing is an effective strategy for containing infectious diseases (Figure 4A).

The diagnostic process in clinical laboratories is designed to inform treatment decisions for individual patients. Therefore, specimen collection by health professionals and individualized testing are prioritized over high capacity and rapid turnaround times. In contrast, efforts to contain infectious disease outbreaks emphasize high capacity and rapid turnaround times for large-scale testing. This workflow utilizes self-sampling, batch testing, and in-house diagnostics. Since clinical diagnostics and monitoring are incompatible, their processes must be kept separate. Furthermore, large-scale testing is only viable when the price of a test is low. Because opportunity costs drive resource allocation in the private sector, large-scale testing is unlikely to be undertaken without external funding. The response model reported here overcomes the limitations of clinical laboratories. We demonstrate how effective monitoring can be achieved by reallocating resources from non medical, nonprofit laboratories, and situate this approach within the framework of infectious disease response strategies.

The effectiveness of monitoring depends on how quickly the test can be developed and whether capacity can be cost efficiently scaled up. qPCR diagnostics are ideal for rapid development because of their modular design. Efficiency is measured by the product cost per specimen when capacity is maximized. Product costs consist of fixed components, like facility rental, and variable components, like reagents. Fixed costs decline per specimen as capacity rises, while variable costs remain constant per specimen. Variable costs dominate when processing specimens individually, so no economies of scale are realized (Figure S5A,B). Batch testing lowers the variable as well as the fixed costs per specimen (Figure S5C, Table S7, S8). Variable costs per specimen decrease as fewer reagents are needed when testing multiple specimens in a single reaction, and as this increases capacity the fixed costs per specimen also decrease. Economical purchasing and in-house production of reagents can further reduce the variable costs, though in-house production requires expertise to ensure the highest quality and delays implementation. The maximum batch size is limited by the test’s sensitivity and the transmissibility of the circulating variant. As cases rise more batches require deconvolution, increasing material use and reducing throughput (Figure S5D). This decrease in capacity and increase in costs can be alleviated by reducing the batch size when the incidence rate increases (Figure S5E-I). Nationwide scaling is most effective when centralized coordination of information systems and diagnostic standards is paired with decentralized specimen processing. This approach reduces turnaround times while local oversight empowers leaders, strengthening accountability, and improving execution.

Public health interventions rely on information gathered through monitoring. Adding anonymized metadata, such as age, gender, location, and virus variant, improves disease modeling and helps target measures where it is most needed. Variant monitoring is especially important, as emerging variants may spread faster requiring stronger measures. At the VBC, a high-throughput sequencing platform was developed for variant detection [3, 38]. This enabled the first detection of the Alpha variant in Austria, which showed ∼50% increase in transmissibility [34, 39]. The information produced by local monitoring is also of global value. Through the GISAID platform, millions of viral genome sequences are shared worldwide, enabling public health responses, such as when and how to update vaccines and diagnostic tests [19, 20]. We quantified mutations relative to the reference genome, which identifies variable regions by recording selected and recurrent mutations at a site cumulatively. While global analysis is biased toward the most frequently sequenced strains in countries with greater resources, stable regions in the genome are less affected by this bias and are reliable for assay design.

Protecting vulnerable groups is a highly effective strategy to save lives [40]. At the start of the pandemic, caregivers faced unprecedented challenges, requiring immediate solutions. Demographic data showed that older people were disproportionately affected [41]. In addition, the communal structure of care homes increases transmission risk. Caritas, a Catholic social aid organization, initiated a simulation study to identify the best strategy to prevent outbreaks in care homes [42]. This study showed that, due to its higher sensitivity, qPCR testing is more effective at reducing outbreaks than rapid antigen testing. Additionally, monitoring twice weekly with same-day results reduces the risk of an outbreak for the Alpha variant. Caritas isolated residents, so that the caregivers became the main route of virus introduction, making immediate monitoring of caregivers critical. This required affordable qPCR tests, supplies and data systems. Additionally, a loop-mediated isothermal amplification point-of-care test was developed for situations requiring an immediate diagnosis [43]. Our partnership with Caritas started with monitoring 240 caregivers in one home and expanded to 17 homes with 2475 caregivers. The VBC provided testing and materials, but de-emphasized specimen collection and contact tracing in order to focus its resources on its core competencies and save time. Collaborating with Caritas, a partner with complementary expertise, medical know-how, commitment and excellent infrastructure, enabled the rapid implementation of monitoring. Similar outcomes for governments, organizations and companies depend on tailored solutions, cross-functional collaboration with the relevant expertise of their partners, and commitment up to the highest organizational level.

The International Health Regulations (IHR) require countries to develop core capabilities for outbreak detection, response and timely reporting. Although the IHR provides a pragmatic legal framework, compliance is limited and the WHO lacks enforcement power. To strengthen implementation, the Global Health Security Agenda introduced a security-based approach, but global systems for outbreak response still lack effective coordination [44]. In many low-income countries, weak public health and clinical infrastructure make outbreak containment unreliable. The COVID pandemic revealed that high-income countries also lack the core capabilities for responding to fast spreading infectious diseases (Figure 4B). This resulted in a disease control only paradigm that undermines public trust, whereas monitoring requires broad public support. Medical care is essential both for trust and disease control. Because health care providers are a primary vulnerable group [40], they must be monitored and have access to personal protective equipment to prevent health system collapse. However, advanced biomedical technologies are ineffective without adequate resources, as demonstrated by the supply chain disruptions in the COVID pandemic (Figure 4B). Stockpiling critical supplies and preparing laboratories, clinics and care homes through regular functional and full-scale exercises is vital for an effective response [45]. Risks can be further reduced by focusing on general principles of prevention, education, communication, accountability, and investment in research frameworks that accelerate knowledge transfer (Table S9). Vaccination, in particular, protects communities against infectious disease outbreaks. Prevalent vaccine hesitancy increases the risk of re-emerging infectious diseases.

The pandemic proved that widespread infections are inevitable without proper prevention and response strategies. In today’s highly connected world, a direct flight between any two cities is shorter than the incubation period of any known human pathogen [46, 47]. State security therefore requires investment in robust regional and global health systems supported by long-term prevention and response strategies.

## Materials & Methods

### Mutation Map and Assay Development

A SARS-CoV-2 assay was established based on the viral genome targets published by the US CDC and Charité. We chose the Luna Probe One-Step RT-qPCR Kit (NEB, #E3007). In this kit the reverse transcription and qPCR are combined. Clinical specimens were used for performance testing. The need for a variant-inclusive assay became apparent when a new variant emerged that evaded detection. Figure S1C shows the work breakdown structure of qPCR assay development. To identify regions with a low mutation rate in the viral genome, a mutation map was generated based on 4,295,664 high-coverage, complete sequences downloaded from GISAID. These sequences were aligned to the reference genome (Wuhan-Hu-1/2019, NCBI RefSeq: NC_045512.2) using unimap v0.1-r41 (https://github.com/lh3/unimap) with parameters: -- MD -a -cxasm5 --cs -t8 [48]. Mutations were quantified at each position in the reference genome using the pysam module (https://github.com/pysam-developers/pysam), excluding bases representing low confidence base calls. Fourteen ∼100 bp stable regions were identified with a cumulative mutation rate between 1% and 20% of the uniform mutation distribution across the viral genome. Ninety primer-probe sets targeting these regions were designed with Primer3 software (https://gear-genomics.com). A text-based search was used to count perfect matches to validate for inclusive specificity. To avoid the high cost of TaqMan probes, primer sets were tested with RT-qPCR using 1 μl/reaction intercalating dye EvaGreen (Fc 1.25 μM, Biotium, #31000). Exclusive specificity was assessed using purified throat wash from healthy individuals, with and without DNaseI digestion. PCR products analyzed on 2.5% agarose gels indicated that most primer sets amplified off-target sequences. RT-qPCR with EvaGreen was performed on throat wash spiked with 25 copies/ μl of synthetic virus (AccuPlex, SARS-CoV-2 Verification Panel, Full Genome, #0505-0168) to validate the absence of primer-dimers. Eight primer sets were selected and tested in combination with TaqMan probes conjugated to the fluorophore FAM. One probe failed to detect SARS-CoV-2 and re-synthesis did not improve results. Multiplexing assays reduce costs, but incompatible assay combinations reduce PCR efficiency. Triplex combinations consisting of two assays targeting different viral genome regions and an oligo-limited human control were evaluated. Fluorophores with distinct emission wavelengths were applied. The primary endpoints of the best performing triplex were assessed as described in the Clinical *Performance Study* section.

### Specimen Collection

A quality management system was implemented in accordance with the WHO handbook based on ISO 15189 [49]. After providing consent for data handling and for procedures in the event of a positive test result, campus employees were tested anonymously. VBC employees signed a consent form authorizing the medical director to access their test results. Self-sampling of specimens was introduced to reduce costs and improve willingness with regular testing. Participants were instructed to collect specimens before breakfast and before brushing their teeth, ensuring sufficient cell yield and avoiding food debris. Specimens were collected by rinsing the back of the throat (oropharynx) with 0.9% NaCl solution (Braun, #3505270), kept in motion by exhaling for 5×5 seconds (gargling) [23]. The solution was then expelled into a 50 ml tube (Falcon, #352070) and 1 ml was transferred with a disposable pipette to a 1.4 ml tube with an external screw cap (Micronic, #MP52761-Y20). On campus, all employees deposited their specimens in a refrigerator located at a central outdoor collection point. For Caritas caregivers, more advanced logistics were required because care homes are located throughout Vienna and batch testing was implemented.

### Specimen Preparation

After specimens arrived in the laboratory, the QR codes of the Micronic tubes (#MP52761-Y20) were scanned using the FluidX Perception HD Whole Rack Reader (Azenta, #20-4018), and the tube rack was disinfected with 70% ethanol. High-throughput, error-free pipetting and reproducibility were achieved by automating the workflow with the IntelliXcap 96 tube capper/decapper (Azenta, #46-8012), the ViaFlo96 liquid handler (Integra, #6001, #6106), and the KingFisher magnetic particle processor (ThermoFisher, #5400630). Wide-bore tips (Integra, #6635) were used to prevent clogging. Specimens exhibit substantial viscosity heterogeneity, which impedes automated handling. To facilitate liquid handling and reduce the risk of cross-contamination, 15 μl of 2M 1,4-Dithiothreitol (DTT, Roth #6908.4) was mixed with each specimen by pipetting, followed by a 10 minute incubation at room temperature (RT). Alternatively, the cheaper sodium ascorbate (Fc 62.5 mM, Roth, #3149.2) can be used for sputolysis. Since sodium ascorbate is a certified food additive (E301), it can also be added directly to the 0.9% NaCl solution, further shortening turnaround time. Both sputolysis methods cause a detection delay of approximately one cycle. To increase test sensitivity, specimens were cell-enriched by centrifugation of 96-tube racks at 500 rcf for 1 minute (Heraeus Multifuge 4 KR, Sorvall rotor 75006475). Subsequently, 800 μl of the supernatant was removed, and the pellet was suspended in the remaining 200 μl. From this suspension, 100 μl was used for nucleic acid extraction, and the remainder was stored at 4°C.

### Nucleic Acid Extraction

The nucleic acid extraction protocol developed by Boom et al. was adapted for automation using carboxylated beads and the KingFisher magnetic bead processor [50]. The DNaseI digestion, which removes genomic DNA, was omitted to reduce costs and processing time. Testing with the CDC-N2 and VBC primer-probe sets showed that this omission did not significantly reduce RNA yield and sensitivity. Prior to lysis, 8 μl of 2M DTT was mixed with 342 μl of Lysis Buffer (80 mM Tris pH 6.4, 5.7 M guanidinium isothiocyanate - Carl Roth, #0017.2, 35 mM EDTA, 2% Triton X-100) and pipetted into each well of a deep-well plate (Ritter, #43001-0502). The liquid handler transferred 100 μl of each cell-enriched specimen into 350 μl Lysis Buffer per well, followed by vigorous mixing for 1 minute by the KingFisher and incubation for 10 minutes at RT to complete lysis. Next, 400 μl ethanol premixed with 20 nl carboxylated magnetic beads (Cytiva, #65152105050450) was added to each well. The KingFisher mixed the lysed specimens with the beads for 5 minutes at RT (mixing comb Ritter, #43001-0503), followed by one wash for 1 minute in 900 μl wash buffer I (4.7 M guanidine HCl, 10 mM Tris pH 6.6, 60% ethanol) and two washes for 30 seconds each in 700 μl wash buffer II (20mM NaCl, 2mM Tris pH 7.5, 80% ethanol). Residual ethanol was removed from the beads by air drying for 3 minutes, and nucleic acids were eluted in 50 μl RNase-free water by incubating the beads at 60°C for 5 minutes in low-profile plates (Ritter, #43001-0501).

### Inactivation of Crude Specimens

An RNase and virus inactivation protocol originally developed for specimens containing Zika or dengue [24] was validated for SARS-CoV-2. Validation is required because the viral envelopes differ between these viruses. The protocol employs a combination of the reducing agent tris(2-carboxyethyl)phosphine (TCEP), the chelating agent ethylenediamine tetraacetic acid (EDTA), and heat. SARS-CoV-2 was detected at an earlier quantification cycle when EDTA was omitted. Therefore, the inactivation protocol was simplified as follows: 18 μl of crude specimen was mixed with 2 μl of 10× TCEP (25mM, pH 7.0) in a low-profile PCR plate, sealed, and incubated at 95°C for 5 minutes in a PCR machine. The heated lid prevents condensation in the caps avoiding incomplete inactivation. TCEP must be pH neutral, as PCR efficiency decreases under acidic conditions. It can be purchased at pH 7.0 (Sigma-Aldrich, #646547) or as a powder (Roth, #HN95.2), which is dissolved in water and adjusted to pH 7.0 with ammonium hydroxide (Sigma-Aldrich, #221228-1L-M).

### Reverse Transcription Quantitative PCR

The Luna Probe One-Step RT-qPCR Kit was used for diagnostic analysis (NEB, no ROX, #E3007). This kit reduces turnaround time and lowers the risk of contamination by combining the reverse transcription (RT) and polymerization in a single step. In addition to assay quality, the guaranteed delivery of large quantities was an important factor in selecting this kit. For high specificity we used TaqMan hydrolysis probes [51], which specifically bind to the target amplicon and act as fluorescent reporters. The TaqMan probe is conjugated with a fluorophore at the 5′ end and a quencher at the 3′ end. During strand synthesis, the 5’-3’ exonuclease activity of Taq polymerase degrades the probe, separating the fluorophore from the quencher. To enable detection of three independent targets in a single reaction, TaqMan probes binding to different targets were conjugated with fluorophores that emit at distinct wavelengths: 6-Carboxyfluorescein (FAM), 5’-Hexachloro-Fluorescein (HEX) and 6-Carboxy-X-Rhodamine (ROX). FAM and HEX were quenched with BMN-Q535, while ROX was quenched with BMN-Q620. The maximum probe length was limited to 27 nucleotides, as background fluorescence increases with probe length. Longer probes can be quenched with multiple quenchers, but this increases the cost.

The master mix for a single reaction:

- 10.00 μl Luna One-Step reaction mix (NEB)
- 1.00 μl Luna WarmStart enzyme mix (RT, NEB)
- 0.10 μl Forward primer (Fc 500nM, Sigma-Aldrich)
- 0.10 μl Reverse primer (Fc 500nM, Sigma-Aldrich)
- 0.05 μl TaqMan probe (Fc 250 nM, biomers.net)
- 1.75 μl RNase-free ddH_2_O (NEB)

For multiplex reactions, additional primers and probes were added and the water was reduced accordingly. A 13 μl master mix was dispensed into each well of a 96-well low-profile PCR plate (Bio-Rad, #HSP9601, #MSB1001) in a clean room using a multi-dispenser. Then, 7 μl of throat wash extract was added to each well using a multichannel pipette in a PCR workstation located in a low-copy room, followed by plate sealing, vortexing and brief centrifugation. Amplification was performed in a CFX96 Touch Real-Time PCR System (Bio-Rad, #1855196) in the high copy room.

The thermocycling program:

- Reverse transcription at 55°C for 10 minutes
- Polymerase activation at 95°C for 2 minutes

45 cycles:

- Denaturation at 95°C for 10 seconds
- Annealing and extension at 57°C for 30 seconds

For direct RT-qPCR with inactivated crude specimens, the same thermocycling program was applied, but the specimen volume was reduced to 4 μl with the water volume adjusted accordingly.

### Controls

A positive human internal control is required to confirm both specimen quality and the integrity of the reaction components. The CDC-RPP30 control assay targets a single exon (1) of the human Ribonuclease P gene, and therefore amplifies both RNA and genomic DNA [52]. This control should not be used when the sample contains genomic DNA. Several intron-spanning human assays were evaluated to identify a control assay that only detects human mRNA. To remove genomic DNA, the nucleic acids bound to the beads were washed with 700 μl wash buffer II, air dried for 1 minute, and then incubated at 37°C for 15 minutes with 100U RNase-free DNaseI (5 μl premixed with 195 μl 1× DNaseI buffer, NEB, #M0303L). The RNA was then purified as described. To obtain extracts that also contain genomic DNA, the DNaseI digestion was omitted. Half of each extract was digested with 350U Ribonuclease A (5 μl of 1 mg/ml, Sigma-Aldrich, #R6513) at 37°C for 40 minutes to remove all RNA. To prevent residual RNA amplification in the RNase-treated sample during RT-qPCR, the reverse transcriptase was inactivated at 95°C for 5 minutes. RT-qPCR with samples that only contain genomic DNA showed detection by the RPP30 assay but not by the B2M assay targeting human β-2-Microglobulin mRNA (Table S1). RT-qPCR with samples that contain RNA, with or without genomic DNA, showed detection by both assays. The CDC-N2-B2M and VBC-O1b-S-B2M multiplexes were optimized by limiting primer (Fc 100nM) and probe (Fc 50nM, ROX-labeled) concentration of the B2M assay, as its target is abundant in the oropharynx. The PCR efficiency of VBC-B2M in multiplex reactions with specimens was determined using LinRegPCR software (Table S4, Figure S4Y). The VBC-B2M assay is reproducible and equally efficient in combination with CDC-N2 and VBC-O1b-S (Table S4). It shows a two-cycle difference between RT-qPCR performed with purified and with inactivated crude specimens, both when combined with CDC-N2 and VBC-O1b-S assays (Figure S4X). Each plate also includes a positive control (synthetic SARS-CoV-2 RNA, Twist, #102019) and a negative control (RNase free water). The positive control must yield amplification, and the negative control must not, before evaluation of the test results can proceed. If the internal control fails to detect human B2M mRNA, the test result is inconclusive and the specimen must be retested. If the retest is also inconclusive, a new specimen must be collected. The results are reported as *positive* if at least one of the two SARS-CoV-2 assays show an exponential fluorescence curve, and as *not* detected if only background fluorescence is observed.

### Clinical Performance Study

The nucleic acid amplification test was developed using one-step multiplex RT-qPCR technology to detect SARS-CoV-2 RNA qualitatively in human throat wash. This test was used in house to monitor employees and inform epidemiological decision making. Conserved regions of the ORF1ab and S genes were used as primer and probe target sites (Figure 2). We evaluated the parameters of the SARS-CoV-2 NAATs in a clinical performance study (CPS). The CPS design was non-interventional, non-randomized, retrospective, open-label and single-laboratory. It was approved by the Ethics Committee of the Vienna Hospitals of the Vinzenz Group (#EK57/2020) and conducted in accordance with the Declaration of Helsinki, Law 14/2007 on Biomedical Research, and ISO 13485 standards. The primary endpoints of the CPS was the evaluation of specificity and sensitivity [31]. Inclusive specificity was determined in silico using high-coverage complete viral genome sequences from 4,295,664 patients worldwide (Table 1, S3). Exclusive specificity was evaluated with throat wash from healthy individuals and by comparison with SARS-CoV, the Sarbecovirus closely related to SARS-CoV-2 (79.5% sequence identity) (Table S2). Analytical sensitivity was determined by replicate RT-qPCR using nucleic acid extracts from throat wash from healthy individuals spiked with synthetic virus (AccuPlex, SARS-CoV-2 Verification Panel, Full Genome #0505-0168) at final concentrations of 0.05, 0.10, 0.20, and 0.40 copy/μl. The binary outcomes of 20 technical replicates at each dilution were fitted with probit regression, and analytical sensitivity was interpolated at 95% (Figure 3A, Figure S4W) [33]. Accuracy was defined as agreement with a standard, and benchmarked against the CDC-N2 reference [22]. We did not use the same kit as the CDC and included an internal control (VBC-B2M). The remaining specimens from 48 patients with confirmed SARS-CoV-2 infection were analysed. Two positive specimens were excluded because they were not detected by the CDC-N2 assay in direct RT-qPCR. All RT-qPCRs performed with specimens from healthy individuals were negative. Specimens were collected and processed as described in the previous sections. Reactions were performed on the same plate and with the same instrument. The quantification cycle (C_q_) threshold was set to the same relative fluorescence units for all reactions. A standard curve measures PCR efficiency under optimal conditions, and requires a broad-range standard that may not be available in an epidemic. Therefore, actual efficiency was calculated for each specimen using the raw fluorescence curve (without baseline subtraction) in the LinRegPCR software (https://gear-genomics.com) [35]. For statistical analysis initial copy numbers were calculated because C_q_ values are exponential.

The amplification reaction follows the equation:

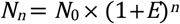

where *N*_n_ = amplicon number, *N*_0_ = initial copy number, *E* = efficiency, and *n* = quantification cycle.

The equation for relative initial copy numbers:

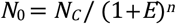

based on *N*_0_ = 100 of the standard, *N*_35_ = *N*_C_ (*C* = constant). A correction was applied for the smaller input volume in direct RT-qPCR. The assays showed a linear dynamic range of 6Log_10_ (CDC-N2: R^2^=0.978, *P*=8.17×10^-40^, VBC-O1b: R^2^=0.919, *P*=9.52×10^-27^, VBC-S: R^2^=0.861, *P*=2.29×10^-21^). Accuracy was assessed by calculating the coefficient of determination (R^2^) with regression analysis (Figure 3C,D, S4E-N). The VBC assays demonstrated higher mean individual efficiencies, but also a wider confidence interval compared to the reference, with higher individual efficiency for low copy number specimens (Figure S4).

### Statistics

Analytical sensitivity follows the Poisson distribution, and is defined as the concentration of viral genomes in a specimen that has a 95% probability of yielding a positive result. This concentration was determined using probit regression, a maximum likelihood ratio estimator that models binary outcomes [33]. Reproducibility was assessed by calculating the coefficient of variation (CV) from 24 replicate reactions performed over three days. The coefficient of determination (R^2^) indicates the proportion of shared variance between assays, indicating the accuracy measured against a standard assay. The distribution of relative initial copy numbers of the clinical specimens was tested using the Shapiro-Wilk test. For normally distributed dependent data, the paired t-test was applied. For non-normally distributed dependent data, the non-parametric paired Wilcoxon signed-rank test was applied. The significance level was set at α=0.05.

### Laboratory Information Management System

As part of the automated workflow, a database application to accelerate specimen handling, minimize error, and deliver test results directly to participants was developed. The application is capable of handling large data sets and separates test results strictly from personal data. The sprint Gantt for LIMS development is shown in Figure S1B. Specimens were managed by uploading their IDs and test results to the backend, with test results published to the frontend. At the backend, incoming specimens were registered by scanning the QR codes on the tubes. Each specimen was stored in the database and assigned to a position in an 8×12 array layout that was maintained throughout the workflow. Each plate received a unique ID, and the layout was exported to a plate export folder to enable error-free upload of specimen codes into the CFX Maestro qPCR software. Once RT-qPCR was completed, the data were exported from the CFX Maestro software into a results folder and uploaded to the database. The application automatically assigned each specimen a *positive* or a *not detected* result based on the presence or absence of C_q_ values. Before publication to the frontend, all results were verified by a human. Each status transition of a specimen in the workflow was tracked with a timestamp. Campus employees were issued an anonymous user ID and password, enabling secure access to the frontend. Test results were retrieved by logging into the website and entering the specimen ID.

### Batch testing

Pooling was outsourced and performed manually to:

1. Conserve critical consumables for the robotic system
2. Reduce laboratory processing time
3. Lower overall costs

Each Caritas employee received a kit containing 12 tests. All tests within a kit were labeled with the same unique barcode. To ensure label durability, barcodes were printed on adhesive white polypropylene film with silicone paper backing, using a Toshiba B-EX4T1 industrial printer (thermal transfer foil: Resin Black/N/OUT, size: 10 mm × 250 mm).

A single test consists of:

- 5.0 ml 0.9% NaCl solution
- Barcoded 5.0 ml safe-lock tube (pooling)
- Barcoded 1.5 ml safe-lock tube (backup)
- Disposable Pasteur pipette
- Separate barcode label

Participants were instructed to divide each specimen between two tubes: one for pooling and one for backup. Each tube was placed in a separate box containing a plastic zipper bag. On the specimen submission list, representing a single pool, participants recorded their name, the date and affixed the corresponding barcode label. Once up to 10 participants had submitted their specimens, the zipper bags and list were replaced. For each pool, a coded Micronic tube was added to the zipper bag containing the 5.0 ml tubes. The human-readable code of the Micronic tube was recorded on the specimen submission list and on both zipper bags. To link all data, the individual specimen codes and the pool code were scanned and connected to the corresponding names in the Caritas database. After the submission deadline, all zipper bags were delivered to the pooling room. In the pooling room, equal amounts (2 ml) of up to 10 specimens were manually poured into a 50 ml tube (Falcon, #352070). To reduce viscosity, 1/20 volume of 1.25M sodium ascorbate was added to the pool. After vortexing 1 ml was transferred to the 1.4 ml Micronic tube. The pooled batch was then processed as described in the previous sections, with the exception that the sputolysis (DTT) step during specimen preparation was omitted. A two-stage batch testing approach was applied: pooled samples were tested first, followed by individual testing of backup samples from positive batches. Backup samples were stored at 4°C. If a pool tested positive, these crude samples were inactivated as described, and directly analyzed with RT-qPCR. After quality control, the medical director was informed of the results. Caritas employees received their results through the Caritas database. In the event of a positive test result, isolation, contact tracing, and retesting by a clinical laboratory were initiated immediately. The test results were submitted to the department for public health.

## Data Availability

All data produced in the present work are contained in the manuscript

## Ethical approval

This study involving human specimens was reviewed and approved by the Ethics Committee of the Vienna Hospitals of the Vinzenz Group (#EK57/2020).

## Conflict of interest

The authors declare that there is no conflict of interest.

## Acknowledgements

We gratefully acknowledge all data contributors for generating the genetic sequence and metadata and sharing via the GISAID Initiative, on which part of this study is based (gisaid.org/EPI_SET_220330me). We thank Raoul Lavaulx-Vrecourt for assistance with estimating fixed costs, Sabina Kula, Kristina Uzunova and Boril Bochev for technical advice, Andreas Untergasser for advice on the individual PCR efficiency, Robert Fritsche-Polanz, Petra Pjevac, Buck Hanson, and Mihaela Peycheva for support. Figure 1 was partially created using icons from BioRender. MPSD and MJK were supported by the Vienna Science and Technology Fund (WWTF) through project COV20-031 given to JZ. The IMP receives funding from Boehringer Ingelheim GmbH and the Austrian Research Promotion Agency (HQ grant FFG-852936).

## Author contributions

Conceptualization: JZ, SW, TW-T, PS, HS, TM, RH, MPSD. Implementation: SW, MW, CU, JT, KS, SR, MA-R, TM, MM, MJK, MR-H, RH, DH, MF, MPSD, NB. Clinical study: VG, MPSD. Data analysis: TN, MPSD with input from JZ. Development of data analysis tools: TN. Development of data management tools: TM. Writing: MPSD with feedback from JZ, ZC, RH and TW-T.

## Vienna COVID-19 Detection Initiative

Stefan Ameres, Institute of Molecular Biotechnology of the Austrian Academy of Sciences (IMBA), Vienna, Austria; Benedikt Bauer, Research Institute of Molecular Pathology (IMP), Vienna, Austria; Nikolaus Beer, IMP, IMBA, and Gregor Mendel Institute (GMI), Vienna, Austria; Katharina Bergauer, IMP, Vienna, Austria; Wolfgang Binder, Max Perutz Labs (MPL), Medical University of Vienna, Vienna, Austria; Claudia Blaukopf, IMBA, Vienna, Austria; Boril Bochev, IMP, IMBA, and GMI, Vienna, Austria; Julius Brennecke, IMBA, Vienna, Austria; Selina Brinnich, Vienna Biocenter Core Facilities (VBCF), Vienna, Austria; Aleksandra Bundalo, IMP, Vienna, Austria; Meinrad Busslinger, IMP, Vienna, Austria; Aleksandr Bykov, IMP, Vienna, Austria; Tim Clausen, IMP, and Medical University of Vienna, Vienna, Austria; Luisa Cochella, IMP, Vienna, Austria; Geert de Vries, IMBA, Vienna, Austria; Marcus Dekens, IMP, Vienna, Austria; David Drechsel, IMP, Vienna, Austria; Zuzana Dzupinkova, IMP, IMBA, and GMI, Vienna, Austria; Michaela Eckmann-Mader, VBCF, Vienna, Austria; Ulrich Elling, IMBA, Vienna, Austria; Michaela Fellner, IMP, Vienna, Austria; Thomas Fellner, VBCF, Vienna, Austria; Laura Fin, IMP, Vienna, Austria; Bianca Valeria Gapp, IMBA, Vienna, Austria; Gerlinde Grabmann, VBCF, Vienna, Austria; Irina Grishkovskaya, IMP, Vienna, Austria; Astrid Hagelkruys, IMBA, Vienna, Austria; Bence Hajdusits, IMP, Vienna, Austria; Buck Hanson, Centre for Microbiology and Environmental Systems Science, University of Vienna, Vienna, Austria; David Haselbach, IMP, Vienna, Austria; Robert Heinen, IMP, IMBA, and GMI, Vienna, Austria; Louisa Hill, IMP, Vienna, Austria; David Hoffmann, IMBA, Vienna, Austria; Stefanie Horer, IMP, Vienna, Austria; Harald Isemann, IMP, Vienna, Austria; Robert Kalis, IMP, Vienna, Austria; Max Kellner, IMP and IMBA, Vienna, Austria; Juliane Kley, IMP, Vienna, Austria; Thomas Köcher, VBCF, Vienna, Austria; Alwin Köhler, MPL, Medical University of Vienna, Vienna, Austria; Darja Kordic, IMP, Vienna, Austria; Christian Krauditsch, IMBA, Vienna, Austria; Sabina Kula, IMP, IMBA, and GMI, Vienna, Austria; Richard Latham, IMP, Vienna, Austria; Marie-Christin Leitner, IMBA, Vienna, Austria; Thomas Leonard, MPL, Medical University of Vienna, Vienna, Austria; Dominik Lindenhofer, IMBA, Vienna, Austria; Raphael Arthur Manzenreither, IMBA, Vienna, Austria; Karl Mechtler, IMP, Vienna, Austria; Anton Meinhart, IMP, Vienna, Austria; Stefan Mereiter, IMBA, Vienna, Austria; Thomas Micheler, VBCF, Vienna, Austria; Paul Moeseneder, IMBA, Vienna, Austria; Tobias Neumann, IMP, Vienna, Austria; Simon Nimpf, IMP, Vienna, Austria; Magnus Nordborg, GMI and IMBA, Vienna, Austria; Egon Ogris, MPL, Medical University of Vienna, Vienna, Austria; Michaela Pagani, IMP, Vienna, Austria; Andrea Pauli, IMP, Vienna, Austria; Jan-Michael Peters, IMP, Medical University of Vienna, Vienna, Austria; Petra Pjevac, Centre for Microbiology and Environmental Systems Science, and Joint Microbiome Facility of the University of Vienna and Medical University of Vienna, Vienna, Austria; Clemens Plaschka, IMP, Vienna, Austria; Martina Rath, IMP, Vienna, Austria; Daniel Reumann, IMBA, Vienna, Austria; Sarah Rieser, IMP, Vienna, Austria; Marianne Rocha-Hasler, Centre for Microbiology and Environmental Systems Science, University of Vienna, Vienna, Austria; Alan Rodriguez, IMP and IMBA, Vienna, Austria; James Julian Ross, IMBA, Vienna, Austria; Harald Scheuch, IMP, IMBA, and GMI, Vienna, Austria; Karina Schindler, IMP, Vienna, Austria; Clara Schmidt, IMBA, Vienna, Austria; Hannes Schmidt, Centre for Microbiology and Environmental Systems Science, University of Vienna, Vienna, Austria; Jakob Schnabl, IMBA, Vienna, Austria; Stefan Schüchner, MPL, Medical University of Vienna, Vienna, Austria; Tanja Schwickert, IMP, Vienna, Austria; Andreas Sommer, VBCF, Vienna, Austria; Johannes Stadlmann, Institute of Biochemistry, University of Natural Resources and Life Sciences (BOKU), Vienna, Austria; Alexander Stark, IMP, Vienna, Austria, and Medical University of Vienna, Vienna, Austria; Peter Steinlein, IMP, IMBA, and GMI, Vienna, Austria; Simon Strobl, VBCF, Vienna, Austria; Qiong Sun, IMP, Vienna, Austria; Wen Tang, IMP, Vienna, Austria; Linda Trübestein, MPL, Medical University of Vienna, Vienna, Austria; Christian Umkehrer, IMP, Vienna, Austria; Sandor Urmosi-Incze, VBCF, Vienna, Austria; Kristina Uzunova, IMP, IMBA, and GMI, Vienna, Austria; Gijs Versteeg, MPL, University of Vienna, Vienna, Austria; Alexander Vogt, VBCF, Vienna, Austria; Vivien Vogt, IMP, Vienna, Austria; Michael Wagner, Centre for Microbiology and Environmental Systems Science, and Joint Microbiome Facility of the University of Vienna and Medical University of Vienna, Vienna, Austria; Martina Weißenböck, IMP, Vienna, Austria; Barbara Werner, VBCF, Vienna, Austria; Ramesh Yelagandula, IMBA, Vienna, Austria; Johannes Zuber, IMP, and Medical University of Vienna, Vienna, Austria.

## Notes

### Competing Interest Statement

The authors have declared no competing interest.

### Funding Statement

Marcus P. S. Dekens and Max J. Kellner were supported by the Vienna Science and Technology Fund through project COV20-031 to Johannes Zuber. The Institute of Molecular Pathology receives funding from Boehringer Ingelheim GmbH and the Austrian Research Promotion Agency (Headquarter grant FFG-852936).

### Summary of Updates

As a description of the model without supporting evidence of its effectiveness would leave an important gap, we have now included a comparison of the incidence rates among monitored caregivers with those of the general population of Vienna (who were not systematically monitored and not vaccinated during the same time frame). These data are presented in Figure 4A. A final paragraph has been added to the Results section describing the analysis. These data further strengthen the evidence for the effectiveness of our early response model. The Summary has been updated, the first two paragraphs of the Discussion section have been refined, and Figure S1A has been refined.

