## Supplemental Figures and Tables for "Early response model for containing newly emerging infectious diseases"

Figure S1

### A Gantt chart for the implementation of infectious disease monitoring

| Code | Task | Predecessor | | Duration (d) |
|---|---|---|---|---|
| **1.1** | **PROJECT MANAGEMENT** | | | |
| 1.1.1 | Project start | | | 1 |
| 1.1.2 | Project coordination | | | 58 |
| 1.1.3 | Project controlling | | | 53 |
| 1.1.4 | Project close down | | | 2 |
| 1.1.5 | **Project accepted – M7** | | | |
| **1.2** | **CREATE CONCEPTS** | | | |
| 1.2.1 | Create overall concept: QMS | | | 7 |
| 1.2.2 | Create concept for RT-qPCR assay | 1.2.1 | SS | 3 |
| 1.2.3 | Contact clinic for specimens | 1.2.1 | SS | 1 |
| 1.2.4 | Create concept for workflow and scale up | 1.2.1 | SS | 3 |
| 1.2.5 | Create concept for information system (LIMS) | 1.2.4 | SS | 5 |
| 1.2.6 | Repurpose or rent facility | 1.2.1 | SS | 10 |
| 1.2.7 | Apply for clinical performance study approval | 1.2.2 | SS | 21 |
| 1.2.8 | Place orders with suppliers | 1.2.1 | SS | 5 |
| 1.2.9 | Recruit staff | 1.2.1 | SS | 20 |
| 1.2.10 | Public authorization of facility | 1.2.6 | FS | 15 |
| 1.2.11 | **Contracts & authorizations concluded – M1** | 1.2.10 | | |
| **1.3** | **DEVELOP INFORMATION SYSTEM** | | | |
| 1.3.1 | Implement backend LIMS | 1.2.5 | FS | 25 |
| 1.3.2 | Implement frontend LIMS | 1.3.1 | SS | 20 |
| 1.3.3 | Test and optimize LIMS | 1.3.2 | FS | 5 |
| 1.3.4 | **LIMS operational – M2** | 1.3.3 | | |
| **1.4** | **DEVELOP ASSAYS AND WORKFLOW** | | | |
| 1.4.1 | Develop RT-qPCR assays | 1.2.2 | SS | 21 |
| 1.4.2 | Clinical performance study | 1.2.7 & 1.4.1 | FS | 5 |
| 1.4.3 | **RT-qPCR assays passed tests – M3** | 1.4.2 | | |
| 1.4.4 | Simplify, optimize, accelerate supply system | 1.2.8 | SS | 24 |
| 1.4.5 | Document standard operating procedure (SOP) | 1.2.4 | FS | 10 |
| 1.4.6 | Implement and optimize small-scale workflow | 1.4.2 & 1.4.4 | FS | 10 |
| 1.4.7 | **Small-scale workflow optimized – M4** | 1.4.6 | | |
| **1.5** | **IMPLEMENT WORKFLOW SCALE UP** | | | |
| 1.5.1 | Install waste disposal systems | 1.2.10 | SS | 2 |
| 1.5.2 | Assemble laboratory automation system | 1.2.6 | FS | 26 |
| 1.5.3 | Simplify and optimize large-scale workflow | 1.5.2 | FS | 10 |
| 1.5.4 | Simplify, optimize and accelerate batch testing | 1.4.6 | FS | 10 |
| 1.5.5 | **Scale up workflow completed – M5** | 1.5.4 | | |
| 1.5.6 | Train laboratory technicians | 1.5.3 | FS | 10 |
| 1.5.7 | Perform external quality test | 1.5.3 | FS | 10 |
| 1.5.8 | **Monitoring laboratory operational – M6** | 1.5.7 | | |

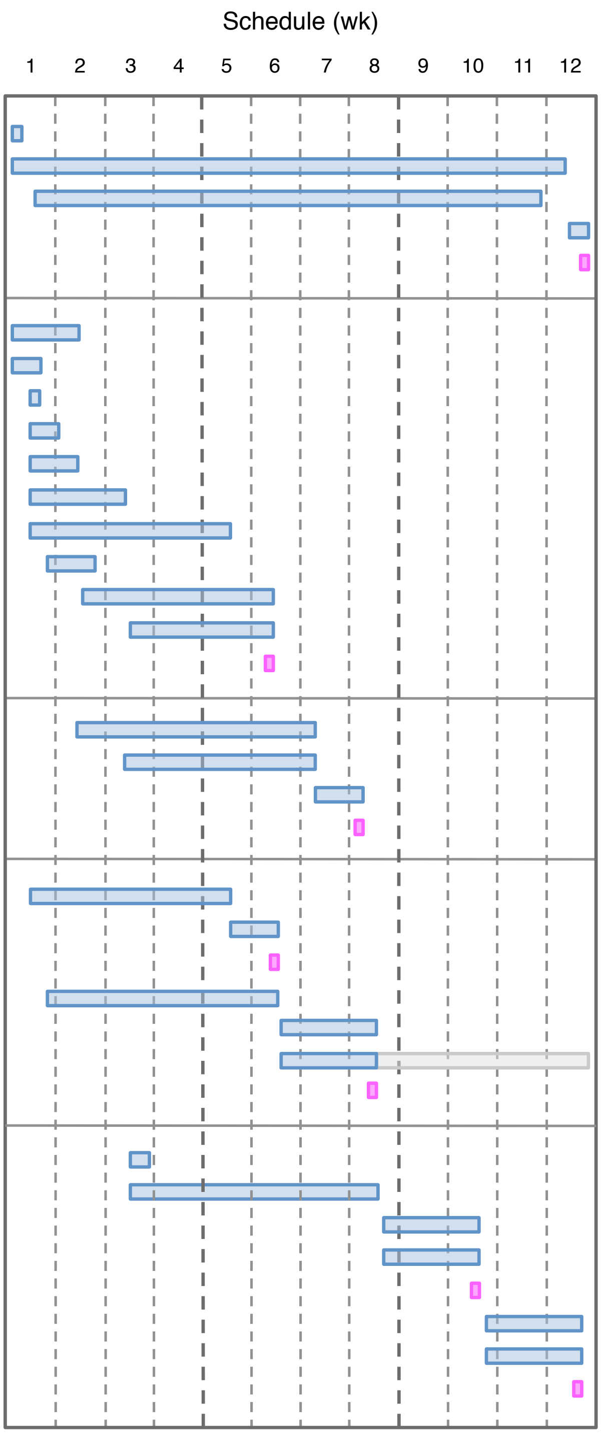

#### B Sprint Gantt for the laboratory information management system

| Code | Tasks | Predecessor | Duration |
|---|---|---|---|
| **2.2** | **SPRINT 0: CREATE CONCEPT** | | |
| 2.2.1 | Define technology stack | - | 2.0d (20h) |
| 2.2.2 | Define UI design | - | 5.0d (60h) |
| 2.2.3 | Set up development configuration | 2.2.1 | 1.0d (10h) |
| 2.2.4 | Sandbox configuration and database setup | 2.2.3 | 2.0d (20h) |
| 2.2.5 | **Concept and configurations concluded – M1** | 2.2.2, 4 | |
| **2.3** | **SPRINT 1: BASIC BACKEND FEATURES** | | |
| 2.3.1 | Analyze current data sources | - | 1.5d (20h) |
| 2.3.2 | Define data structure | 2.3.1 | 2.0d (25h) |
| 2.3.3 | Define data formats | 2.3.1 | 0.5d (5h) |
| 2.3.4 | Specify barcodes and label design | 2.3.2 | 1.5d (20h) |
| 2.3.5 | Implement user management | - | 3.0d (30h) |
| 2.3.6 | Implement permission and role management | - | 3.0d (30h) |
| 2.3.7 | **Basic backend functions done – M2** | 2.3.4, 5, 6 | |
| **2.4** | **SPRINT 2: SAMPLE MANAGEMENT** | | |
| 2.4.1 | General and data definition of sample management | - | 1.0d (10h) |
| 2.4.2 | Implement “sample view” feature | 2.4.1 | 2.0d (30h) |
| 2.4.3 | Implement “sample search” feature | 2.4.2 | 1.5d (20h) |
| 2.4.4 | Implement sample CRUD operations | 2.4.2 | 2.0d (20h) |
| 2.4.5 | Implement sample import / export functionality | 2.4.2 | 2.0d (20h) |
| 2.4.6 | Implement frontend UI | - | 5.0d (40h) |
| 2.4.7 | **Sample management functions done – M3** | 2.4.6 | |
| **2.5** | **SPRINT 3: ANALYSIS MANAGEMENT** | | |
| 2.5.1 | General and data definition | - | 1.0d (10h) |
| 2.5.2 | Define and implement sample workflow and status history | 2.5.1 | 1.0d (15h) |
| 2.5.3 | Define analysis control sets | 2.5.1 | 1.0d (10h) |
| 2.5.4 | Implement analysis verification | 2.5.1 | 3.0d (50h) |
| 2.5.5 | Implement analysis import / export functionality | 2.5.4 | 1.0d (20h) |
| 2.5.6 | Implement frontend UI | - | 5.0d (40h) |
| 2.5.7 | **Sample analysis functions done – M4** | 2.5.5, 6 | |
| **2.6** | **SPRINT 4: PLATE MANAGEMENT** | | |
| 2.6.1 | Define and implement plate workflow and status | - | 0.5d (5h) |
| 2.6.2 | Implement “plate view” feature | 2.6.1 | 3.0d (50h) |
| 2.6.3 | Implement “plate search” feature | 2.6.2 | 1.0d (15h) |
| 2.6.4 | Implement plate CRUD operations | 2.6.2 | 1.5d (15h) |
| 2.6.5 | Implement plate import / export functionality | 2.6.2 | 0.5d (5h) |
| 2.6.6 | Implement “batch” feature | 2.6.1 | 2.0d (20h) |
| 2.6.7 | Implement frontend UI | - | 5.0d (40h) |
| 2.6.8 | **Plate management functions done – M5** | 2.6.2, 3, 7 | |
| **2.7** | **SPRINT 5: LIMS INTEGRATION** | | |
| 2.7.1 | Sample and plate integration label printer | - | 3.0d (30h) |
| 2.7.2 | Sample integration code scanner | - | 2.0d (20h) |
| 2.7.3 | Implement IT backup strategy | - | 2.0d (10h) |
| 2.7.4 | Design and develop API | - | 2.5d (20h) |
| 2.7.5 | Performance optimization | - | 4.0d (30h) |
| 2.7.6 | Implement email features | - | 1.5d (10h) |
| 2.7.7 | Implement frontend UI | - | 5.0d (40h) |
| 2.7.8 | **LIMS integration done – M6** | 2.7.1, 2, 7 | |
| **2.8** | **SPRINT 6: TEST & IMPROVE** | | |
| 2.8.1 | Test and improve user permission management | - | 2.0d (20h) |
| 2.8.2 | Test and improve sample management | - | 2.0d (20h) |
| 2.8.3 | Test and improve plate management | - | 2.0d (20h) |
| 2.8.4 | Test and improve frontend UI | - | 2.0d (20h) |
| 2.8.5 | Setup and test production infrastructure | - | 2.0d (20h) |
| 2.8.6 | Execute workload and performance tests | 2.8.5 | 1.0d (20h) |
| 2.8.7 | Perform final tests | 2.8.6 | 2.0d (20h) |
| 2.8.8 | **Testing done – M7** | 2.8.7 | |
| 2.8.9 | **LIMS operational – M8** | 2.8.8 | |

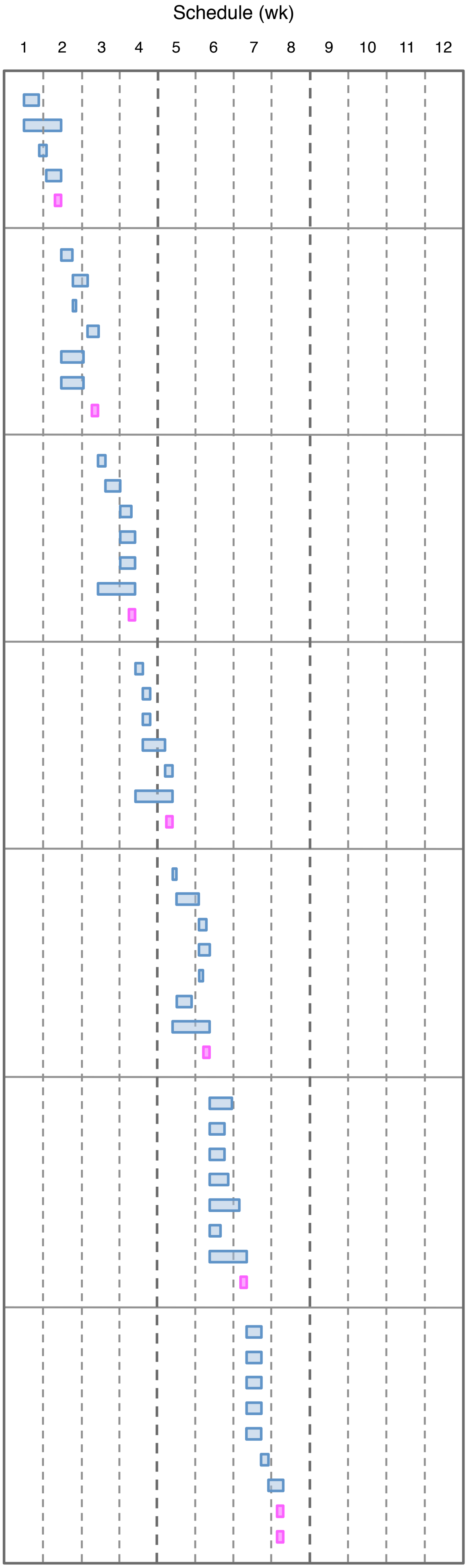

#### C Gantt chart for qPCR assay development

| Code | Task | Predecessor | | Duration (d) |
|---|---|---|---|---|
| **3.2** | **CREATE CONCEPT** | | | |
| 3.2.1 | Create concept for RT-qPCR assay | | | 3 |
| 3.2.2 | Contact clinic for specimens | | | 1 |
| 3.2.3 | Apply for clinical performance study approval | 3.2.1 | SS | 21 |
| 3.2.4 | **Concept and approval concluded – M1** | 3.2.3 | | |
| **3.3** | **IN SILICO ASSAY DESIGN** | | | |
| 3.3.1 | Download pathogen genome sequences | | | 1 |
| 3.3.2 | Align sequences and quantify mutations | 3.3.1 | FS | 12 |
| 3.3.3 | Design assays that target stable regions | 3.3.2 | FS | 1 |
| 3.3.4 | Validate inclusive specificity of assays | 3.3.3 | FS | 0.2 |
| 3.3.5 | **Variant-inclusive assays designed – M2** | 3.3.4 | | |
| 3.3.6 | Iterative assay validation: repeat 3.3.1 & 2 & 4 | 3.3.4 | FS | 13 |
| 3.3.7 | **Variant-inclusive assays retained** | 3.3.6 | | |
| **3.4** | **EMPIRICAL ASSAY SELECTION** | | | |
| 3.4.1 | Collect pathogen-negative specimens | 3.2.1 | FS | 13 |
| 3.4.2 | Establish pathogen detection protocol | 3.2.1 | FS | 13 |
| 3.4.3 | Order oligonucleotides | 3.3.3 | SS | 1 |
| 3.4.4 | Select human assay for exclusive specificity | 3.4.3 | FS | 2 |
| 3.4.5 | Select pathogen primers for exclusive specificity | 3.4.3 | FS | 3 |
| 3.4.6 | Validate absence of primer-dimers | 3.4.3 | FS | 0.1 |
| 3.4.7 | Combine primers with probe and validate assays | 3.4.5 | FS | 1 |
| 3.4.8 | Combine assays, optimize and select multiplex | 3.4.7 | FS | 3 |
| 3.4.9 | Determine analytical sensitivity with standard | 3.4.8 | SS | 3 |
| 3.4.10 | Determine reproducibility (CV) | 3.4.8 | SS | 3 |
| 3.4.11 | **Specific sensitive multiplex developed – M3** | 3.4.10 | | |
| **3.5** | **CLINICAL PERFORMANCE STUDY** | | | |
| 3.5.1 | Collect clinical specimens | 3.2.3 | FS | 5 |
| 3.5.2 | Determine $C_q$ and individual PCR efficiency | 3.2.3 | FS | 5 |
| 3.5.3 | Statistics on copy numbers versus reference assay | 3.5.2 | SS | 1 |
| 3.5.4 | Determine shared variance with reference assay | 3.5.2 | SS | 0.2 |
| 3.5.5 | **RT-qPCR assays passed tests – M4** | 3.5.4 | | |
| 3.5.6 | **In-house in vitro diagnostic operational – M5** | | | |

Schedule (wk)

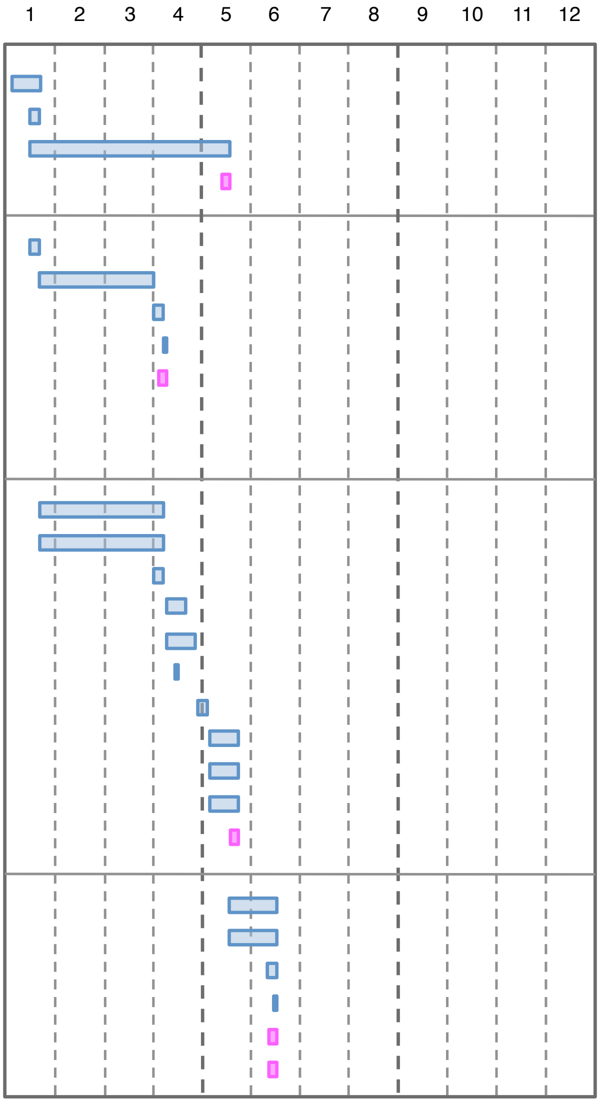


#### D Organogram for the implementation of infectious disease monitoring

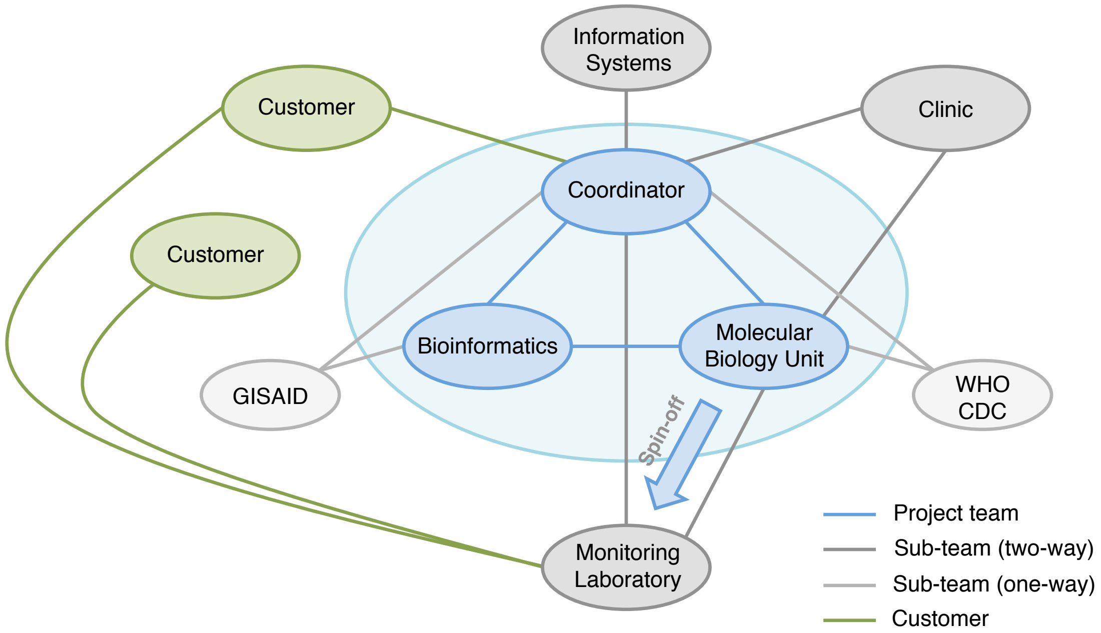

**Figure S1.** Framework for the implementation of infectious disease monitoring

A. Gantt chart for the implementation of infectious disease monitoring. This plan assumes no prior preparations and that ordered supplies arrive without the extended delays typical of a pandemic. The left section shows the work breakdown structure, with tasks listed in order of completion. The right section shows the schedule, where the duration of each task is indicated by blue bars. After completion of the pilot, the prototype workflow continues with specimen testing until the automated monitoring laboratory becomes fully operational (extension of bar shown in grey). Milestones (purple bars) indicate the achievement of key objectives. Dependencies illustrate which tasks can be performed concurrently and which require the completion of a predecessor. Abbreviations: SS = Start to Start, FS = Finish to Start, QMS = Quality Management System, LIMS = Laboratory Information Management System.

B. Sprint Gantt for the laboratory information management system (LIMS). This sub-project is included in the "Gantt chart for the implementation of infectious disease monitoring" under codes 1.2.5 and 1.3. Each task was performed by at least two software developers from a team of four. Abbreviations: API = Application Programming Interface, CRUD = Create Read Update Delete, UI = User Interface.

C. Gantt chart for qPCR assay development. This sub-project is included in the "Gantt chart for the implementation of infectious disease monitoring". A bar is not displayed for the iterative assay validation (code 3.3.6), as this task was carried out outside the shown time frame. Abbreviations: CV = coefficient of variation, $R^2$ = coefficient of determination, IVD = in vitro diagnostics.

D. Organogram for the implementation of infectious disease monitoring. The organizational network structure promotes adaptability by emphasizing communication, collaboration, and resource sharing. Blue circles represent the project team, which interacts with external partners and the customer (grey and green circles). The project manager coordinates the monitoring implementation. The molecular biology unit implements published assays and/or develops assays in collaboration with a bioinformatician. Sequences of circulating virus variants are made available through the GISAID platform. The clinic provides patient specimens to validate assay quality. The software development team implements the database application framework. Once the molecular biology unit has developed the assays and piloted the prototype workflow, and the IT team has delivered the LIMS application, the monitoring laboratory is implemented. The monitoring laboratory is spun off to avoid tension between innovation and control. Abbreviations: CDC = Centers for Disease Control and Prevention, GISAID = Global Initiative on Sharing All Influenza Data, WHO = World Health Organization.

Figure S2

Example of a layout for a qPCR monitoring workflow

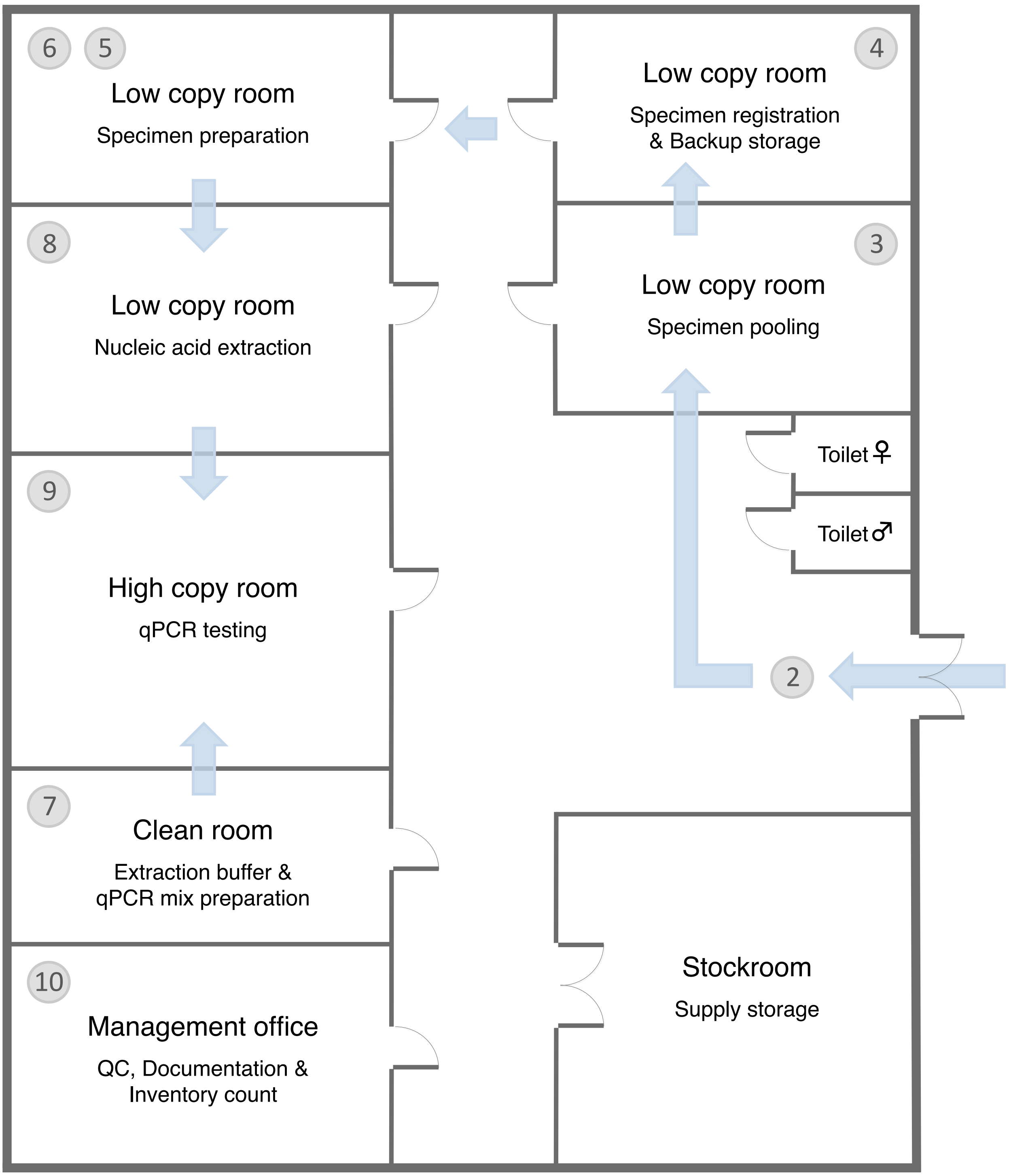

**Figure S2.** Example of a layout for a qPCR-based monitoring workflow

To minimize the risk of amplicon contamination, the diagnostic workflow is divided into separate rooms. The steps described in figure 1 are allocated to the clean room, low copy room, and high copy room, as shown in the floor plan.

Figure S3

Matches and mismatches between assays and SARS-CoV-2 variant pools

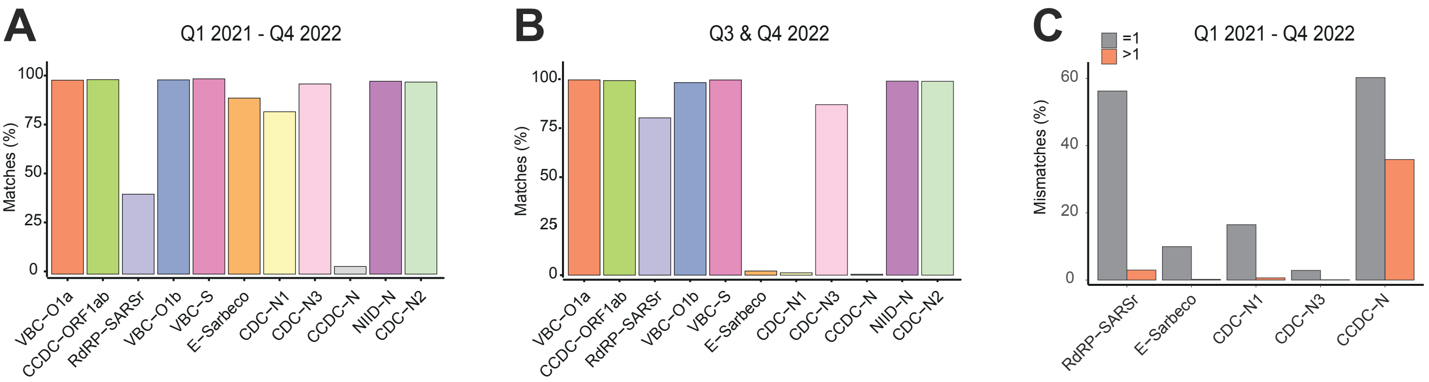

**Figure S3.** Matches and mismatches between assays and SARS-CoV-2 variant pools

A. Match rates of published and variant-inclusive assays based on 4,295,664 analyzed SARS-CoV-2 genomes collected during Q1 2021 - Q4 2022. The RdRP-SARSr, E-Sarbeco, CDC-N1 and CCDC-N assays show reduced inclusive specificity, indicating an increased risk of false negative test results.
B. Match rates of published and variant-inclusive assays based on 42,874 analyzed SARS-CoV-2 genomes collected during Q3 and Q4 2022. Analysis of the most recent part of the time frame shows a pronounced decrease in inclusive specificity for the E-Sarbeco and CDC-N1 assays, but an increase for the RdRP-SARSr assay.
C. Percentage of single and multiple mismatches for assays and their target regions across 4,295,664 analyzed SARS-CoV-2 genomes. With the exception of CCDC-N, assays targeting variable regions in the SARS-CoV-2 genome encounter a single mismatch in the majority of genomes. A single mismatch may have no effect on sensitivity, a minor effect on sensitivity, or give a false negative result. Mismatches at multiple positions within the target sequence further increase the likelihood of false negative test results.

Figure S4 qPCR performance parameters of the CDC and VBC SARS-CoV-2 assays

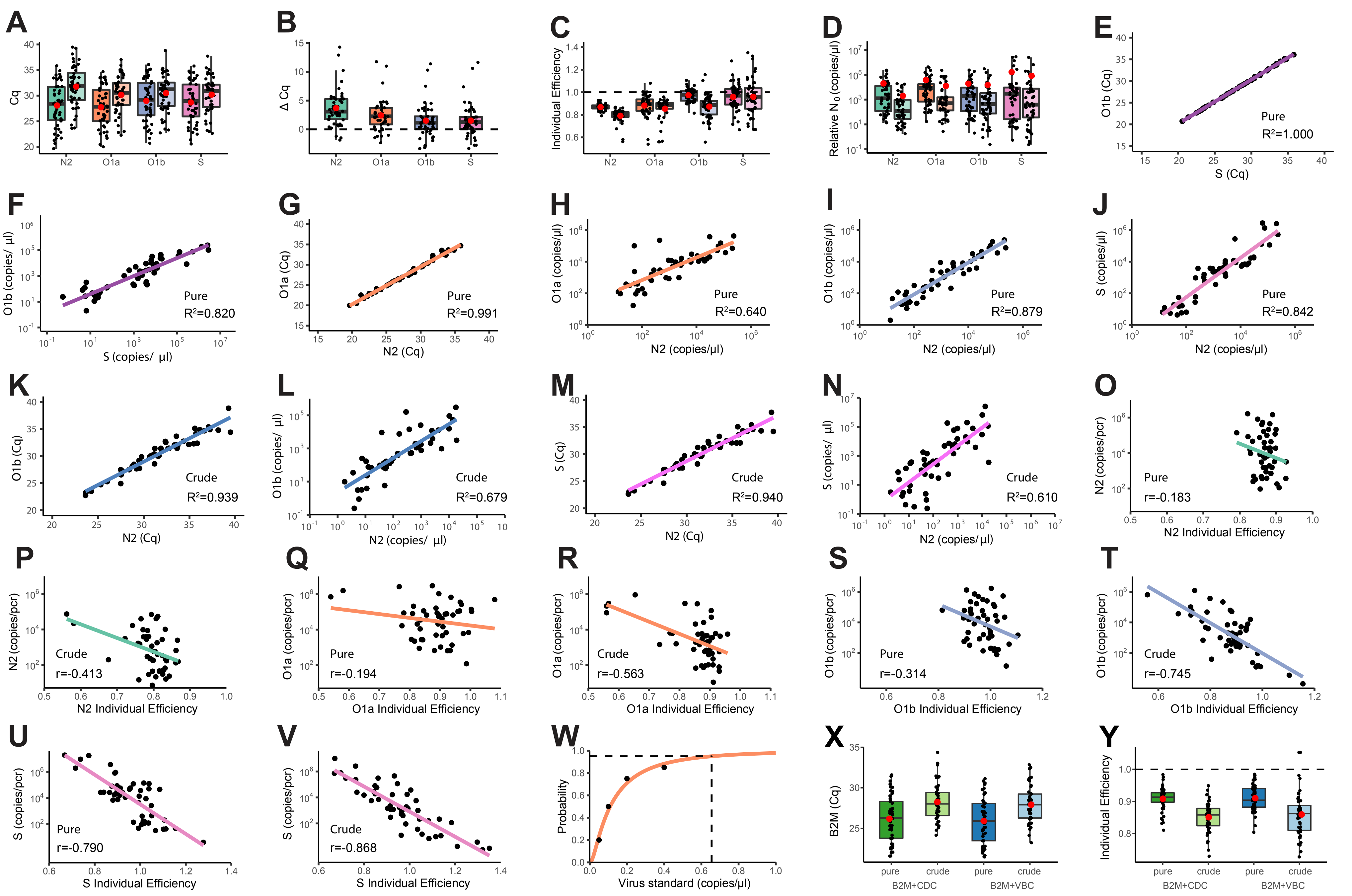

### Figure S4

**A’**

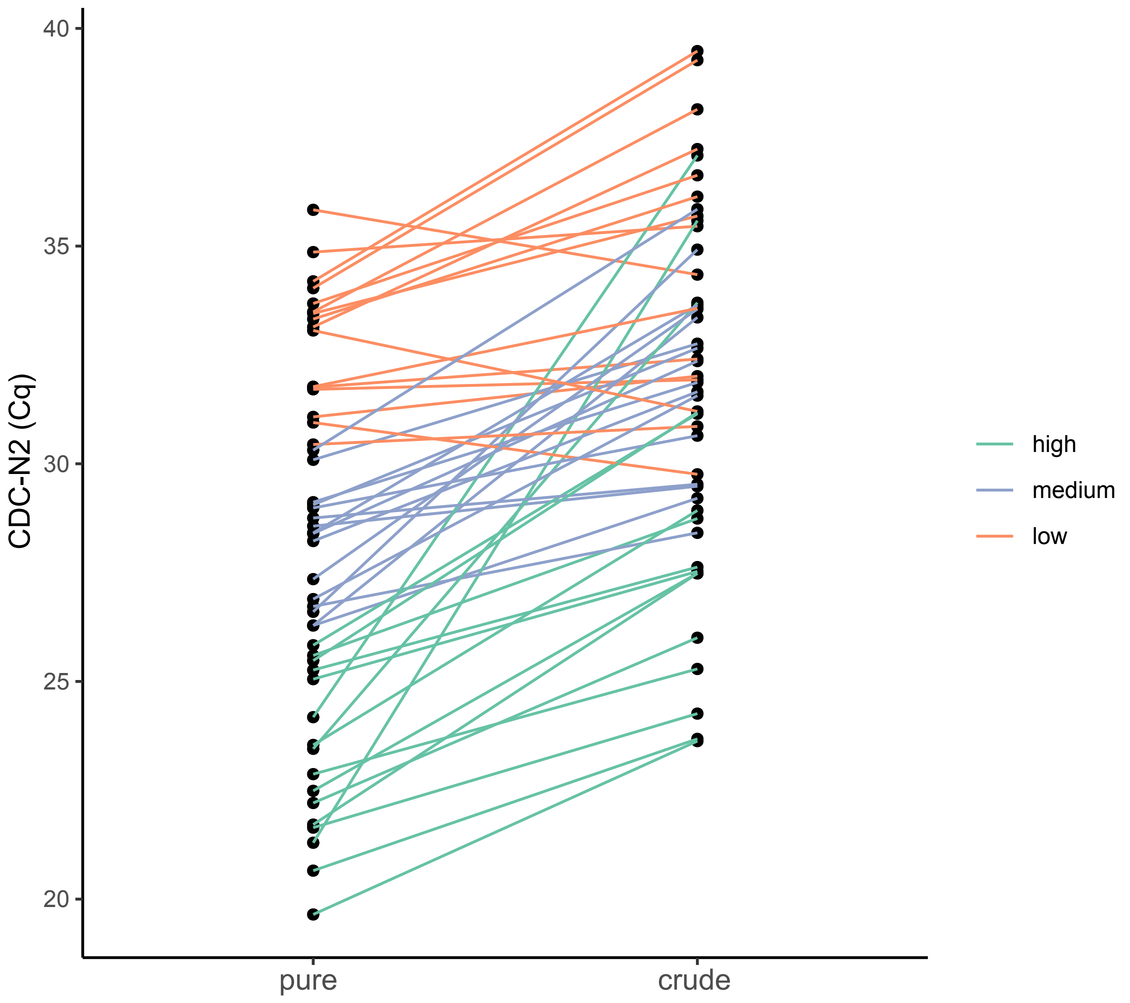

**Figure S4.** qPCR performance parameters of the CDC and VBC SARS-CoV-2 assays

A. Boxplot of $C_q$ values from 48 clinical specimens analyzed with CDC-N2, VBC-O1a, VBC-O1b, and VBC-S assays.

B. $C_q$ differences ($\Delta C_q$) between RT-qPCR with purified or inactivated crude samples for each assay. The $\Delta C_q$ of the CDC-N2 is significantly greater than the $\Delta C_q$ of the VBC-O1a ($P$=9.38×10$^{-9}$), VBC-O1b ($P$=1.68×10$^{-9}$) and VBC-S ($P$=1.66×10$^{-9}$).

C. Individual qPCR efficiencies calculated from fluorescence curves of reactions with each specimen. For qPCR with purified specimens, the CDC-N2 efficiencies are significantly lower than those of the VBC-O1b ($P$=2.37×10$^{-14}$) and VBC-S ($P$=2.13×10$^{-6}$). For RT-qPCR with crude specimens, the CDC-N2 efficiencies are significantly lower than those of the VBC-O1a ($P$=7.0×10$^{-5}$), VBC-O1b ($P$=1.07×10$^{-5}$) and VBC-S ($P$=7.66×10$^{-8}$). A comparison of assay performance between specimen preparation methods shows significant differences for the CDC-N2 ($P$=2.83×10$^{-9}$) and VBC-O1b ($P$=8.45×10$^{-7}$).

D. Relative initial copy number ($N_0$) for each specimen tested across all assays. For RT-qPCR with purified specimens, the VBC-O1b ($P$=0.959) and VBC-S ($P$=0.415) show no significant difference from the CDC-N2 reference, while VBC-O1a ($P$=0.002) shows a significant difference. For RT-qPCR with crude specimens, the VBC-O1a ($P$=3.46×10$^{-5}$), VBC-O1b ($P$=6.39×10$^{-6}$) and VBC-S ($P$=3.65×10$^{-5}$) show significant differences from the CDC-N2 reference. Significant differences between specimen preparation methods were observed for CDC-N2 ($P$=3.44×10$^{-7}$), VBC-O1a ($P$=4.94×10$^{-5}$) and VBC-O1b ($P$=0.020, Z/√n=0.34), with small effect size, but not for VBC-S ($P$=0.079). The coefficients of variation (CV) of the CDC-N2, VBC-O1a, VBC-O1b and VBC-S are as follows: $CV_{pure}$=2.42, $CV_{crude}$=2.24, $CV_{pure}$=2.35, $CV_{crude}$=2.99 , $CV_{pure}$=2.41, $CV_{crude}$=3.33, $CV_{pure}$=3.36, $CV_{crude}$=4.49. The larger CV for VBC-S compared to the reference is likely due to its application in a triplex rather than a duplex reaction. As the polymerase units and extension time are the same for both multiplexes, greater variation in amplification efficiency can be expected in the triplex.

E. Regression of $C_q$ values from VBC-O1b and VBC-S multiplexes. The R-squared ($R^2$=1.000, $P$=3.36×10$^{-78}$) shows that 100% of the variation in purified clinical specimen $C_q$ values is shared.

F. Regression of initial copy numbers ($N_0$) from the VBC-O1b and VBC-S multiplexes. The R-squared ($R^2$=0.820, $P$=9.99×10$^{-19}$) shows that 82% of the variation in purified clinical specimen $N_0$ values is shared.

G. Accuracy of $C_q$ values for VBC-O1a. The R-squared ($R^2$=0.991, $P$=8.27×10$^{-49}$) shows that 99% of the variation in purified clinical specimen $C_q$ values is shared with the reference.

H. Accuracy of $N_0$ values for VBC-O1a. The R-squared ($R^2$=0.640, $P$=9.12×10$^{-12}$) shows that 64% of the variation in purified clinical specimen $N_0$ values is shared with the reference.

I. Accuracy of $N_0$ values for VBC-O1b. The R-squared ($R^2$=0.879, $P$=9.66×10$^{-23}$) shows that 88% of the variation in purified clinical specimen $N_0$ values is shared with the reference.

J. Accuracy of $N_0$ values for VBC-S. The R-squared ($R^2$=0.842, $P$=4.6×10$^{-20}$) shows that 84% of the variation in purified clinical specimen $N_0$ values is shared with the reference.

K. Accuracy of $C_q$ values for VBC-O1b. The R-squared ($R^2$=0.939, $P$=1.52×10$^{-29}$) shows that 94% of the variation in crude clinical specimen $C_q$ values is shared with the reference.

L. Accuracy of $N_0$ values for VBC-O1b. The R-squared ($R^2$=0.679, $P$=6.36×10$^{-13}$) shows that 68% of the variation in crude clinical specimen $N_0$ values is shared with the reference.

M. Accuracy of $C_q$ values for VBC-S. The R-squared ($R^2$=0.940, $P$=1.04×10$^{-29}$) shows that 94% of the variation in crude clinical specimen $C_q$ values is shared with the reference.

N. Accuracy of $N_0$ values for VBC-S. The R-squared ($R^2$=0.610, $P$=5.94×$10^{-11}$) shows that 61% of the variation in crude clinical specimen $N_0$ values is shared with the reference assay.
O. Linear regression of $N_0$ values and individual PCR efficiencies for the purified clinical specimens tested with the reference. The correlation coefficient (r=-0.183) indicates a weak negative relationship.
P. As in O, for CDC-N2 with crude specimens (r=-0.413).
Q. As in O, for VBC-O1a with purified specimens (r=-0.194).
R. As in O, for VBC-O1a with crude specimens (r=-0.563).
S. As in O, for VBC-O1b with purified specimens (r=-0.314).
T. As in O, for VBC-O1b with crude specimens (r=-0.745).
U. As in O, for VBC-S with purified specimens (r=-0.790).
V. As in O, for VBC-S with crude specimens. The correlation coefficient (r=-0.868) indicates a negative relationship, with PCR efficiency increasing as copy number decreases.
W. Probit regression showing the limit of detection for VBC-O1a (0.66 copy/μl).
X. $C_q$ values of 48 clinical specimens tested with the VBC-B2M human assay in different multiplexes. Note: Primer and probe limitation was used for VBC-B2M.
Y. Individual PCR efficiencies derived from the fluorescence curve of the VBC-B2M reaction with each clinical specimen. Note: The same primer and probe limitation was used for VBC-B2M in the different multiplexes.
A'. Connections between the $C_q$ values of the purified sample and the $C_q$ values of the inactivated crude sample of the same specimen, from reactions with CDC-N2 shown in A.

### Figure S5

#### Relation between incidence rate, capacity and estimated cost

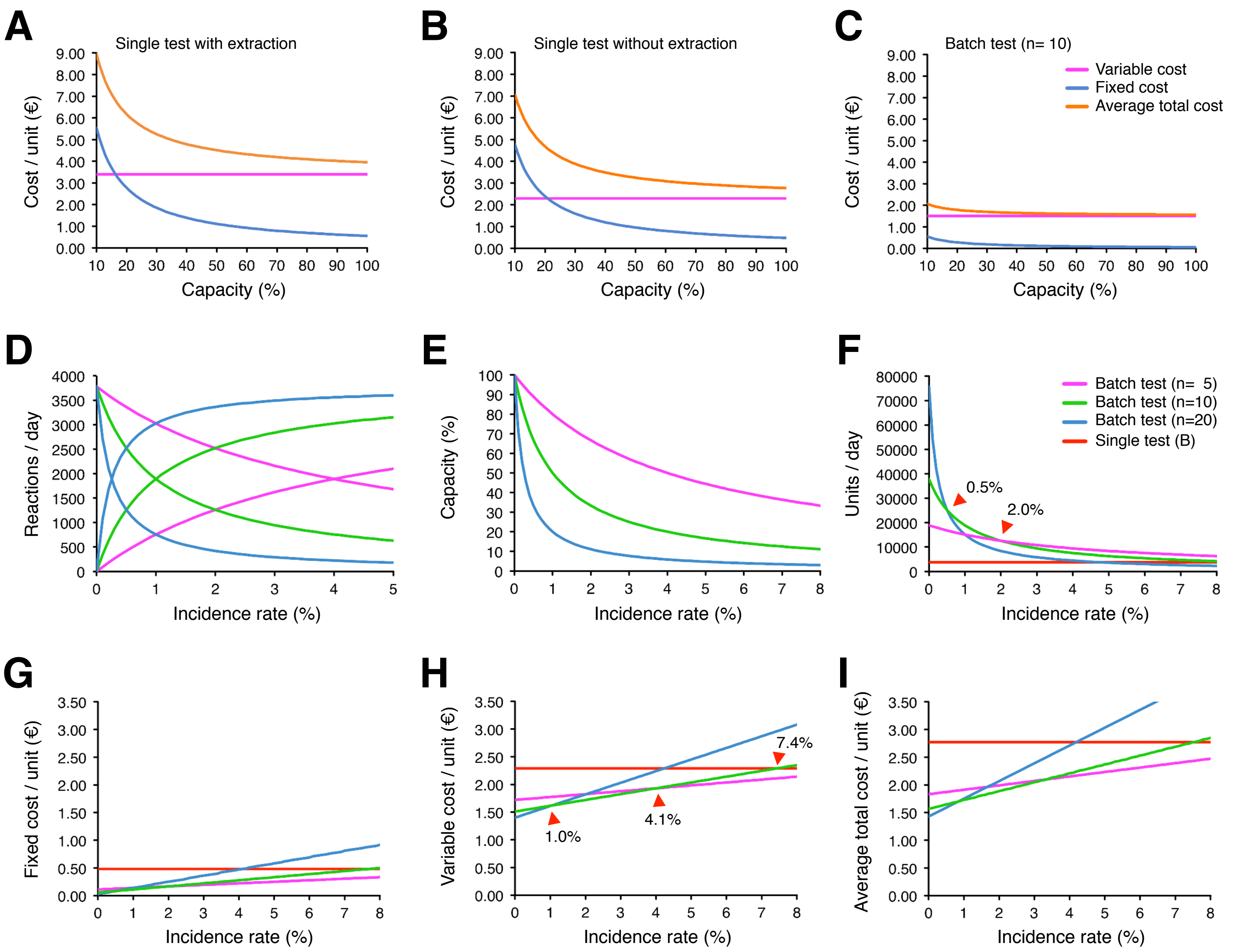

**Figure S5.** Relation between incidence rate, capacity and estimated cost

A. Unit cost of individual testing with specimen purification, showing variable (purple), fixed (blue), and average total costs (orange). Variable costs remain constant, while fixed costs decrease as more specimens are tested.

B. Unit cost of individual testing without specimen purification, using crude specimens in qPCR, which results in a substantial reduction in the variable cost per unit.

C. Unit cost of batch testing at an incidence rate is 0%. In this case, ten specimens are pooled, the pool is purified, and RT-qPCR is performed with the extract. Batch testing reduces fixed costs per unit by increasing capacity, and reduces variable costs per unit by testing multiple specimens in a single reaction. Variable costs can be divided into two components: qPCR reagents and plastics. Switching from single to batch testing significantly reduces reagent costs. However, the cost of plastics remains unchanged, as each person requires a test kit to submit their specimen, regardless of scale.

D. Relationship between batch testing and incidence rate. As incidence increases, fewer batches can be tested because deconvolution (individual testing of backup samples from positive pools) replaces batch reactions. To illustrate the effect across different batches sizes, pools of 5, 10, and 20 specimens are represented by purple, green and blue curves, respectively. Downward-sloping curves represent the number of batch reactions; upward-sloping curves represent the number of deconvolution reactions. For example, pooling 10 specimens instead of 5 yields the same number of batches at 0% incidence, while doubling the number of specimens tested. However, as incidence rises, larger batches generate more deconvolution reactions, leading to greater reductions in overall testing capacity. Because laboratory capacity is fixed, we conservatively assume that positive specimens are distributed across different batches to avoid congestion.

E. Relationship between incidence rate and testing capacity. The maximum number of specimens that can be tested per day is normalized to 100% for each batch size.

F. Relationship between incidence rate and the number of specimens tested per day for each batch size. At an incidence rate of 0.5%, pools of 10 specimens outperform pools of 20; at 2.0%, pools of 5 specimens outperform pools of 10 (red arrowheads).

G. Relationship between incidence rate and fixed cost per unit for each batch size. Fixed cost per unit increases with incidence, as fewer specimens can be tested.

H. Relationship between incidence rate and variable cost per unit for each batch size. Variable costs per unit (materials) increase with incidence, as the number of deconvolution reactions rises. At ≥1% incidence, pools of 10 specimens are more cost-efficient than pools of 20; at ≥4%, pools of 5 are more cost-efficient. To minimize costs, only variable unit costs are considered, as fixed costs are sunk.

I. Relationship between incidence rate and average total cost per unit for each batch size.

Table S1. Oligonucleotides of SARS-CoV-2 and human control qPCR assays[1]

| Assay ID | Forward Primer | Begin | End | Reverse Primer | Begin | End | TaqMan Probe[4] | Begin | End | Amplicon |
|---|---|---|---|---|---|---|---|---|---|---|
| VBC-O1a | ACAACATTGCTTTGATATGGA | 8385 | 8405 | GCAGCACTACGTATTTGTTT | 8463 | 8444 | TCATGTCATTGTCTGAACAACTACGA | 8418 | 8443 | 79 bp |
| VBC-O1b | TCACAGGGCTCAGAATATGACT | 17842 | 17863 | GCCTACTTTTGCTCTGGTAATAGC | 17949 | 17926 | CTCAAACCACTGAAACAGCTCACTCT | 17876 | 17901 | 108 bp |
| VBC-S | GTTGAGGCTGAAGTGCAAAT | 24521 | 24540 | CTGCAGCTCTAATTAATTGTTGAGT | 24611 | 24587 | AGGTTGATCACAGGCAGACTTCAAAGT | 24545 | 24571 | 91 bp |
| CDC-N2[2] | TTACAAACATTGGCCGCAAA | 29164 | 29183 | GCGCGACATTCCGAAGAA | 29230 | 29213 | ACAATTTGCCCCCAGCGCTTCAG | 29188 | 29210 | 67 bp |
| VBC-B2M[3] | GACTTTGTCACAGCCCAAGATAG | Exon2 | Exon2 | CCAAATGCGGCATCTTCAAAC | Exon4 | Exon4 | TGGGATCGAGACATGTAAGCAGCA | Exon2 | Exon3 | 81 bp |

[1] Assay target regions in the SARS-CoV-2 genome are indicated in figure 2 (NCBI ID: NC_045512.2).
[2] Reference assay from the United States Centers for Disease Control and Prevention (Lu et al.).
[3] Control assay: Exon spanning design to detect only human β-2-Microglobulin mRNA (NCBI ID: NM_004048.4).
[4] TaqMan Probe Fluorophores and Quenchers:
The VBC-O1b and VBC-S probes are labeled with either the FAM or HEX fluorophore and BMN-Q535 quencher.
The CDC-N2 probe is labeled with the FAM fluorophore and BMN-Q535 quencher.
The VBC-B2M (human control) probe is labeled with the ROX fluorophore and the BMN-Q620 quencher.
All TaqMan probes were synthesized by Biomers.net GmbH (https://www.biomers.net/).

Table S2. Exclusive specificity: Mismatches between SARS-CoV-2 assays and SARS-CoV[1]

| Assay ID | Mismatches | | | |
|---|---|---|---|---|
| | Forward Primer | Probe | Reverse Primer | Assay |
| VBC-O1a | 7 | 6 | 1 | 14 |
| VBC-O1b | 2 | 3 | 5 | 10 |
| VBC-S | 4 | 3 | 5 | 12 |
| CDC-N2 | 0 | 5 | 2 | 7 |

[1] SARS-CoV BJ01 sequence of virus collected from father of index case (NCBI ID: AY278488.2).

#### Table S3. Inclusive specificity: qPCR assay match rates for SARS-CoV-2 variants of concern[1]

| Assay ID | Match rates for Varants of Concern (%) | | | | |
|---|---|---|---|---|---|
| | B.1.1.7 (Alpha) | B.1.351 (Beta) | B.1.617.2 (Delta) | P.1 (Gamma) | B.1.1.529 (Omicron) |
| VBC-O1a | 99.59 | 99.85 | 99.83 | 98.33 | 99.55 |
| VBC-O1b | 99.77 | 99.66 | 99.75 | 99.54 | 99.84 |
| VBC-S | 99.92 | 99.93 | 99.76 | 99.89 | 99.86 |
| CDC-N1 | 98.12 | 91.35 | 96.85 | 98.98 | 1.44 |
| CDC-N2 | 98.92 | 98.45 | 99.55 | 97.88 | 98.58 |
| CDC-N3 | 98.74 | 98.02 | 95.85 | 98.15 | 82.51 |
| CCDC-N | 0.16 | 1.31 | 0.38 | 5.71 | 0.36 |
| CCDC-ORF1ab | 99.77 | 99.67 | 99.67 | 98.07 | 99.16 |
| E-Sarbeco | 99.58 | 99.82 | 99.67 | 99.66 | 2.89 |
| RdRP-SARSr | 99.72 | 99.81 | 6.37 | 99.94 | 57.37 |

[1] As defined by the WHO. The Phylogenetic Assignment of Named Global Outbreak Lineages and between parentheses the by the WHO assigned "easy-to-say labels for key variants" are shown. Sequences were downloaded from: https://gisaid.org/

#### Table S4. Reproducibility of the CDC and VBC SARS-CoV-2 assays

| | | Duplex qPCR assays[1] | | Triplex qPCR assays[1] | | |
|---|---|---|---|---|---|---|
| | | CDC-N2 | VBC-B2M | VBC-O1b | VBC-S | VBC-B2M |
| Pure specimen[2] | CV of $C_q$ | 0.23 | 0.44 | 0.55 | 0.55 | 0.47 |
| | CV of $N_0$ | 3.52 | 7.52 | 9.44 | 9.63 | 8.19 |
| Crude specimen[2] | CV of $C_q$ | 1.81 | 0.36 | 1.64 | 1.61 | 0.32 |
| | CV of $N_0$ | 37.44 | 6.08 | 43.00 | 40.73 | 5.50 |

[1] All multiplexes include the control assay that detects human β-2-Microglobulin mRNA (B2M).
[2] CV values from 24 technical replicates qPCRs with purified and crude samples of the same specimen.
Abbrs.: CV = Coefficient of Variation, $C_q$ = Cycle quantification, $N_0$ = Initial copy number.

#### Table S5. PCR efficiency of the CDC and VBC SARS-CoV-2 assays

| | | Duplex qPCR assays[1] | | Triplex qPCR assays[1] | | |
|---|---|---|---|---|---|---|
| | | CDC-N2 | VBC-B2M | VBC-O1b | VBC-S | VBC-B2M |
| Pure trial[2] | CI of E (%) | 86.8 ± 0.8 | 90.8 ± 0.8 | 97.4 ± 1.6 | 95.9 ± 3.2 | 91.0 ± 1.2 |
| Crude trial[2] | CI of E (%) | 79.1 ± 1.7 | 85.1 ± 0.9 | 87.3 ± 2.9 | 96.0 ± 9.5 | 85.9 ± 1.5 |

[1] All multiplexes include the control assay that detects human β-2-Microglobulin mRNA (B2M).
[2] 95% confidence interval to estimate the true mean individual PCR efficiency (E) from the assays.
A conserved region of the ORF1a gene was used as the target site for the VBC-O1a assay, in multiplex with VBC-B2M: $E_{pure}$=88.3±2.7%, $E_{crude}$=85.4±2.6% and $CV_{pure}$=5.65 ($N_0$), $CV_{crude}$=38.05 ($N_0$).
Abbrs.: CI = Confidence Interval.

Table S6. Weekly incidence of population and monitored caregivers[1]

| Weeks (Q4 2020 & Q1 2021) | Incidence rate (%) Vienna population[2] | Incidence rate (%) Vienna caregivers |
|---|---|---|
| October, week 42 | 1.2108 | 0.0000 |
| October, week 43 | 0.9960 | 1.3514 |
| October, week 44 | 1.7651 | 1.0965 |
| November, week 45 | 2.3583 | 1.1111 |
| November, week 46 | 2.2886 | 0.6003 |
| November, week 47 * | 3.3433 | 0.6250 |
| November, week 48 * | 2.6648 | 0.3765 |
| December, week 49 * | 1.5007 | 0.3521 |
| December, week 50 | 1.5705 | 0.2191 |
| December, week 51 | 1.2237 | 0.1995 |
| December, week 52 | 0.7415 | 0.3132 |
| December, week 53 * | 0.6125 | 0.2664 |
| January, week 01 * | 0.8931 | 0.8652 |
| January, week 02 * | 0.5261 | 0.8202 |
| January, week 03 * | 0.5767 | 0.0711 |
| January, week 04 | 0.3030 | 0.1332 |
| February, week 05 | 0.3700 | 0.1541 |
| February, week 06 | 0.3632 | 0.0956 |
| February, week 07 | 0.3399 | 0.1773 |
| February, week 08 | 0.6369 | 0.0447 |
| March, week 09 | 0.8898 | 0.0510 |
| March, week 10 | 0.8557 | 0.0785 |
| March, week 11 | 1.4733 | 0.0387 |
| March, week 12 | 1.8284 | 0.1161 |
| March, week 13 | 1.5071 | 0.1638 |
| April, week 14 * | 1.9030 | 0.0816 |
| April, week 15 * | 1.4301 | 0.1163 |
| April, week 16 * | 0.7853 | 0.0779 |
| April, week 17 * | 0.6641 | 0.0775 |
| May, week 18 | 0.2463 | 0.0382 |
| May, week 19 | 0.1936 | 0.0406 |
| May, week 20 | 0.2308 | 0.0000 |
| May, week 21 | 0.1321 | 0.0425 |

[1] Covering the second and third surges during Q4 2020 and Q1 2021.
[2] Data from Bundesministerium für Soziales, Gesundheit, Pflege und Konsumentenschutz.
* Weeks when city-wide hard lockdowns were imposed.

Table S7. Work in process trial accounts for single testing and batch testing[1]

| Process<br>Costs (€/year)[3] | Single qPCR<br>pure specimen | Single qPCR<br>crude specimen | Batch qPCR[2]<br>0 % positive | Batch qPCR[2]<br>74 % positive |
|---|---|---|---|---|
| Material Costs | 3,200,148 | 2,155,394 | 14,212,422 | 2,562,584 |
| Direct Labor[4] | 312,690 | 265,550 | 312,690 | 312,690 |
| Facility Rental | 123,840 | 123,840 | 123,840 | 123,840 |
| Equipment Depreciation[5] | 63,500 | 38,500 | 63,500 | 63,500 |
| Other Costs[6] | 21,750 | 21,750 | 21,750 | 21,750 |
| End Balance (Debit) | 3,721,928 | 2,605,034 | 14,734,202 | 3,084,364 |
| Maximum Capacity | | | | |
| Units (specimens/year) | 941,220 | 941,220 | 9,412,200 | 1,120,500 |

[1] These are product costs at maximum capacity. Fixed costs were estimated and variable costs are for triplex reactions.
[2] Calculated for batches of 10 pooled purified specimens.
[3] Calculated for 249 working days per year and 8 hours per working day.
[4] Estimated labor costs for laboratory manager and 5 biomedical analysts (BMA) in 2021 or 4 for crude specimen processing.
[5] Straight-line depreciation over 10 years, €0 salvage value and €0 disposal costs was applied to all equipment:

2 Integra ViaFlo Liquid Handling Pipettors
3 KingFisher Magnetic Particle Processors
7 Bio-Rad CFX96 PCR Systems
1 Bio-Rad C1000 Thermal Cycler
5 Biological Safety Class II Cabinets
2 IntelliXcap Capper & Decappers
1 FluidX Tubes Rack Reader
1 Heraeus 4KR centrifuge
5 PCR Workstations
1 Agilent Bravo Automated Liquid Handling Platform
Miscellaneous equipment: computers, refrigerators, pipettes etc.

[6] Utilities, waste disposal and cleaning services.

Table S8. Relative cost analysis for single testing and batch testing[1]

| Process<br>Costs (€/unit)[3] | Single qPCR<br>pure specimen | Single qPCR<br>crude specimen | Batch qPCR[2]<br>0 % positive | Batch qPCR[2]<br>74 % positive |
|---|---|---|---|---|
| Material Costs | 3.40 | 2.29 | 1.51 | 2.29 |
| Direct Labor[4] | 0.33 | 0.28 | 0.03 | 0.28 |
| Facility Rental | 0.13 | 0.13 | 0.01 | 0.11 |
| Equipment Depreciation[5] | 0.07 | 0.04 | 0.01 | 0.06 |
| Other Costs[6] | 0.02 | 0.02 | 0.00 | 0.02 |
| Average Per-Unit Cost | 3.95 | 2.77 | 1.57 | 2.75 |

Product costs consist of fixed and variable components. The fixed costs were estimated. The variable component, materials, can be further divided into reagents and plastics. Performing qPCR directly on single crude specimens eliminates the costs of nucleic acid extraction reagents and plastics. Batch testing significantly reduces the qPCR reagents costs. Since each person requires a test kit to submit a specimen, regardless of whether testing is performed individually or in batches, batch testing does not substantially reduce plastics costs. For single and batch testing the reagents costs were further reduced by multiplexing. All costs were rounded to the nearest number. To avoid rounding errors, the exact line-item costs were used to calculate the average unit cost. Price is assumed to be at or above variable cost, and output is assumed to be at maximum capacity. Revenue and margin are not included, as this was a nonprofit initiative. For reference, the price of a qPCR test from a private sector clinical laboratory in Vienna at the end of 2020 was €120 (including 21% Value Added Tax). The maximum capacity is the same for qPCR with purified and crude specimens, as both rely on the same number of qPCR machines and have identical turnaround times.

Table S9. Risk analysis for infectious disease outbreaks

| Risk description | Risk cause | Risk effect | Measure |
|---|---|---|---|
| Monitoring preparedness | Insufficient infrastructure for monitoring | Undocumented infection and rapid transmission of pathogen | Develop capabilities for rapid detection and response |
| No responsibilities[1] | Responsibilities are not assigned | Decision-making by non-experts without accountability | Assign accountable expert(s) |
| Information source[1] | Reliable information source not communicated | Scarce resources are not used efficiently | All leaders must use one reliable information source |
| Budgets[1] | No budgets allocated for emergency measures | Delayed measures or no measures | Assign budgets to specific emergency measures |
| Emergency exercises[2] | No periodic emergency exercises | Weaknesses in response strategy go undetected | Validate strategy by periodic exercises |
| Global air travel | Air travel facilitates transmission of pathogen | Minimizes the time to control transmission of pathogen | Introduce aviation regulation for passenger health monitoring |
| Regulation paralysis | Regulations impede monitoring implementation | Insufficient or inefficient monitoring facilitates rapid transmission of pathogen | Regulation must take diagnostic process for epidemiological applications into account |
| Supply disruptions | High demand during epidemic exceeds manufacturing capacity | Severe delays in the delivery of critical supplies impedes health care and monitoring | Stockpile critical supplies & minimize reliance on global supply chains |
| Political incentives[2] | Public health officials' positions depend on politics | Political instead of medical and epidemiological incentives influence resource allocation | Appoint independent public health officials / experts |
| Inferior diagnostics | Relaxing diagnostic standards during an epidemic | Inferior diagnostics on the market increase outbreak size | Set diagnostic standards for monitoring during epidemic |
| Variants of concern | Variants emerge that evade detection | False negative test results increase outbreak size | Implement variant monitoring and variant-inclusive diagnostics |
| Amplicon contamination | No physical separation between low and high-copy procedures in qPCR workflow | Laboratory shutdown due to amplicon contamination results in decrease in capacity | Separate procedures into clean, low and high-copy rooms & never open PCR plates |
| Fragility of the health system | Insufficient resources to expand health care capacity in an emergency | Inability of patients to access health care & disease control only paradigm reduces trust | Create a robust health system & invest in technology to remotely diagnose and treat patients |
| Vulnerable groups | Vulnerable groups ignored | Increased mortality and burden on health system | Create strategy to protect vulnerable groups |
| Disease model | Sub-par disease model algorithm and data input | Unreliable forecasts result in less effective measures | Invest in disease modeling & include GDPR / HIPAA compliant metadata |
| Delayed information | Lack of real-time data | Decision-making based on delayed information reduces effectiveness of measures | Invest in surveillance systems that provide real-time data |
| Vaccines[2] | Inadequate vaccine supply & sole focus on preventing critical illness and death | Vaccine shortages & poor infection prevention | Expand through second sourcing manufacturing & improve: total protection, temperature stability, one and done. |
| Public hesitancy | Reduced trust in science and government | No cooperation: Higher probability of re-emerging infectious diseases & ineffective outbreak control | Communicate through trusted sources & invest in reliable health system |
| International cooperation | Insufficient international public health coordination | Higher probability of infectious disease outbreaks / epidemics | Invest in knowledge and resource transfer |

[1] Crimson Contagion 2019 functional exercise report: https://www.governmentattic.org/38docs/HHSaarCrimsonContAAR_2020.pdf
[2] Gates B., How to prevent the next pandemic, 2022, New York, Alfred A. Knopf
